# Evidence-based blood tests for monitoring adults with hypertension in primary care: rapid review, routine data analyses, and consensus study

**DOI:** 10.1101/2025.04.01.25325024

**Authors:** Martha MC Elwenspoek, Rachel O’Donnell, Catalina Lopez Manzano, Sarah Dawson, Lewis Buss, Katie Charlwood, Christina Stokes, Francesco Palma, Alastair D Hay, Penny Whiting, Jessica Watson

## Abstract

**Background:** Substantial variation in testing rates in adults with hypertension across UK primary care suggest that patients are not receiving optimal monitoring.

**Aim:** To develop a minimal set of evidence-based blood tests for adults with hypertension.

**Design:** Rapid review, routine data analyses, and consensus study.

**Setting:** Primary care.

**Method:** We developed an initial list of tests which were recommended by guidelines or used commonly. We created filtering questions to examine the rationale of each test. To answer each question, we performed stepwise rapid reviews of evidence cited by guidelines, high-quality studies, systematic reviews, and individual studies. A consensus group, including clinicians and patients, voted to include or exclude each test in the testing panel based on the evidence. If there was no consensus (>80%), additional evidence was sought through rapid reviews or analyses of primary care records, which was subject to further voting.

**Results:** We identified 16 tests that are routinely ordered for people with hypertension. We found consistent and good evidence that eGFR to detect chronic kidney disease and HbA1c to detect diabetes is beneficial for patients. We found no or inconsistent evidence of the benefit of routinely measuring lipids, electrolytes, haemoglobin, thyroid function tests, clotting tests, calcium, ferritin, folate acid, or vitamin B12. We found good evidence that there is no benefit in routinely monitoring liver function, inflammation markers, or brain natriuretic peptide.

**Conclusion:** We identified a minimal set of evidence-based tests that should be offered to adults with hypertension. Implementing these recommendations could reduce harms associated with unwarranted variation in care. Further research is needed to clarify the role of tests with inconsistent evidence and determine the optimal frequency of testing.

**Key messages:** *What is already known on this topic:* There is substantial variation in the use of blood tests to monitor people with hypertension suggesting that many patients are not receiving optimal care. Over-testing can cause patient harm, including patient anxiety, unnecessary downstream tests, referrals, and overdiagnosis, as well as wasting limited NHS resources.

*What this study adds:* We have developed a minimal testing panel for patients with hypertension based on the best available evidence and a consensus process. The panel includes eGFR to screen for chronic kidney disease, HbA1c to screen for diabetes mellitus, potassium for patients on ACE inhibiters and angiotensin II receptor blockers.

*How this study might affect research, practice or policy – summarise the implications of this study:* Unnecessary testing could be prevented if guidelines and local protocols recommended minimal testing sets and made clear additional tests should only be added if clinically indicated. Future research needs to address optimal testing intervals and test thresholds.

## INTRODUCTION

Hypertension is a common long-term condition. Approximately one in four adults in the UK have hypertension.^1^ It is the leading preventable risk factor for cardiovascular disease (CVD) mortality and morbidity, including heart failure, coronary heart disease, stroke, chronic kidney disease, peripheral arterial disease, and vascular dementia.^2^ Strict management and good blood pressure control through medication or lifestyle changes reduce the risk of secondary conditions.^3^ However, only two in five middle-aged people on treatment for hypertension are adequately controlled.^4^

Hypertension monitoring includes periodic measurement of blood pressure, as well as checking blood and urine tests. Optimal monitoring with blood tests can help detect secondary conditions early. However, guidance on monitoring is largely based on expert opinion,^5^ and testing rates vary substantially between GP practices,^6,7^ suggesting that many patients do not receive optimal care. An analysis of routine primary care data and a survey amongst GPs suggest that the majority of patients receive more tests than recommended.^7,8^

Over-testing is not only a waste of limited NHS resources, it can cause patient harm, including patient anxiety, unnecessary downstream tests, referrals, and overdiagnosis.^9^ Overuse of tests also increases GP workload through reviewing additional test results and costs of further investigations.^10^

Therefore, there is a need to develop evidence-based testing strategies that include a minimal set of tests for people with long-term conditions. Standardised testing panels have the potential to ensure that people get the tests that they need and only receive additional tests when there is a clinical indication. Here, we describe the development of a minimal testing panel of blood tests that are necessary for adults with hypertension using rapid reviews, routine data analysis, and a consensus process.

## METHODS

### Identifying candidate tests

Tests were selected that are currently ordered for patients with hypertension in primary care,^7^ that GPs say they would order for the average hypertensive patient,^8^ and/or that are recommended by UK guidelines.^5^

Tests were categorised by reason for testing, either to screen for secondary conditions and/or adverse treatment effects. We only considered drug monitoring for people on stable treatment, and did not consider monitoring during drug titration or treatment initiation. We also only considered the most commonly prescribed first-line antihypertensives: angiotensin-converting enzyme (ACE) inhibitors, Angiotensin II receptor blockers (ARB), thiazide type diuretic (TTD) and calcium channel blockers (CCB), and side effects listed on British National Formulary.

### Filtering questions

We determined a list of filtering questions that needed to be answered with ‘yes’ for a test to be a useful monitoring test. For secondary condition screening, these were: 1) Are people with the long-term condition at an increased risk of developing the secondary condition that the test is able to detect? 2) Is there anything the clinician can do to manage or treat this? 3) Are there clear benefits of earlier detection or treatment? For adverse treatment effects, these were: 1) Are people with the long-term condition at an increased risk of developing the side effect that the test is able to detect? 2) Is there anything the clinician can do to manage or treat this? 3) Are there clear benefits of earlier detection or treatment?

### Rapid review methods

We looked at the following sources for evidence in favour or against each test: 1) NICE guidelines and clinical knowledge summaries (including references), 2) systematic reviews by searching KSR Evidence, 3) primary studies by searching MEDLINE, Embase, and CENTRAL. We only proceeded to the next source if insufficient evidence was found.

Search strategies were developed by an information specialist (SD) (Supplementary Tables S1-5). In short, terms were combined for hypertension, specific tests, secondary conditions, or side effects. Records were screened in two steps using Rayyan^11^: first, titles and abstracts were screened excluding clearly irrelevant records, followed by full text screening of reports considered potentially relevant. For pragmatic reasons, guidance and systematic reviews were screened by two reviewers independently, whereas for primary studies 20% of randomly selected records were double screened. Standardised data extraction forms were developed in Excel and tested by two reviewers and amended where necessary. The inclusion criteria, number of included studies and patients, and summary estimates were extracted from reviews. The study design, population characteristics, number of included patients, and results were extracted from primary studies. Risk of bias (ROB) was assessed using ROBIS for systematic reviews,^12^ RoB 2 for RCTs,^13^ and ROBINS-E for cohort studies.^14^ One reviewer conducted data extraction and risk of bias assessments.

### Consensus process and additional evidence

The consensus group consisted of three patient representatives, four GPs, one primary care nurse practitioner, and one renal consultant. Evidence identified for each candidate test in the rapid reviews was circulated to the group ahead of the meeting (See supplementary files – Evidence report). The quality of evidence was judged as “good” if we found high quality meta-analyses and/or high quality studies with large sample sizes, “moderate” if we found meta-analysis with some quality concerns and/or several primary studies showing similar results but some may have quality concerns, or “weak” if the evidence consisted of a single primary study with a small sample size with or without quality concerns or several single primary studies with conflicting results.

At an in-person meeting, members of the consensus group voted on whether to include or exclude each test from a minimal testing panel for hypertensive patients or whether additional evidence was needed. If no consensus (>80%) was reached, the evidence was discussed followed by a second vote. If still no consensus was reached, the test was selected for additional evidence.

To answer specific questions that arose during the consensus meeting and to address certain gaps in the evidence, additional rapid reviews and incidence analyses using routine primary care data were conducted. The reviews addressed the following questions using the methods described above: “does lipid profile testing or Q-RISK assessment increase the chance of patients following lifestyle advice?” and “What is the incidence of new onset T2DM in a population with hypertension?”. The incidence analyses estimated the incidence of abnormal sodium and potassium levels in people with hypertension who had normal levels at diagnosis and the methods are described here.^15^ In short, we used the clinical practice research datalink (CPRD) to identify people with hypertension diagnosed between 2011-2019 and age-sex- and practice-matched controls. We used Cox regression to analyse time to first abnormal blood result (sodium <135 or >145mEq/L, potassium <3.5 or >5.5 mEq/L), to first clinically significant abnormal result (sodium <130 or >150 mEq/L, potassium <3.0 or > 6.0 mEq/L).

In a second consensus meeting where the group consisted of three patient representatives, five GPs, two primary care nurses, and one renal consultant, the additional evidence was voted on using the process described above. If no consensus was reached, a third vote was held without the option of “additional evidence” where we asked to group to include or exclude the test in absence of further evidence.

### Patient and Public Involvement

Three patient representatives were included in the consensus group with equal voting rights to the clinicians. In addition, two patient representatives (CS and FP) have been involved in this study from inception to completion as active members of the project management group.

## RESULTS

### Candidate tests

We identified 16 tests to consider for a routine tests for people with hypertension. All 16 tests are currently used or recommended to screen for secondary conditions (Table 1), and 4 tests were also considered for detecting adverse treatment effects (Table 2).

**Table 1.** Tests to screen for secondary conditions – answers to filtering questions.

| Test | Reason for testing | Are people with hypertension at risk? | Can you intervene? | Benefits of earlier detection? | Consensus meeting outcome | Offer routinely to everyone with HTN? | Evidence gaps/comments |
| --- | --- | --- | --- | --- | --- | --- | --- |
| **eGFR** | To detect chronic kidney disease | Yes, the risk of kidney disease is 2 to 3-fold higher in those with HTN compared to those without (good evidence). | **Yes** | **Yes** | 88% consensus to include this test. | **YES** | The consensus group agreed that there is a clear benefit to monitoring eGFR in people with HTN. |
| **HbA1C** | To detect type 2 diabetes mellitus (T2DM) | Yes, the prevalence of T2DM is almost 4 times higher in hypertensive patients compared to normotensive patients (good evidence). People with HTN are at an increased risk of new onset T2DM (good evidence). Increases in blood pressure increase the risk of T2DM and decreases in blood pressure reduces the risk of T2DM (good evidence). | **Yes** | **Yes** | 100% consensus to include | **YES** | The evidence supports the rationale to monitor HbA1c to detect T2DM in people with HTN. |
| **Potassium** | To detect hyper- or hypokalaemia | The prevalence of hyperkalaemia (>5.0 mmol/L) is higher in hypertensive patients than in normotensive patients (moderate evidence).<br>We estimated the incidence of abnormal potassium levels in patients with HTN who had normal levels at diagnosis. After taking age, sex, and<br>variation in testing rates into account, we found that clinically significant abnormal potassium levels were NOT more common in HTN than in the control group (weak evidence). |  |  | 100% consensus to exclude | **NO** | This evidence suggests that testing at diagnosis may be sufficient.<br>We acknowledge that in most UK laboratories potassium will be automatically checked alongside sodium and eGFR. |
| **Sodium** | To detect hyper- or hyponatraemia | No published evidence was found on the prevalence of abnormal serum sodium levels in HTN patients compared to the general population.<br>We estimated the incidence of abnormal sodium levels in patients with HTN who had normal levels at diagnosis. After taking age, sex, and variation in testing rates into account, we found that clinically significant abnormal sodium levels were more common in HTN than in the control group (weak evidence). |  |  | **100% consensus to exclude** | **NO** | The incidence analysis does not take into account that many of these people are on medications that can impact sodium levels, which may explain the increased risk of abnormal sodium levels.<br>For people not taking any medication, the consensus group agreed that regular sodium monitoring is not needed.<br>We acknowledge that in most UK laboratories sodium will be automatically checked alongside potassium and eGFR. |
| **Lipid profile** | To detect dyslipidaemia and assess | People with HTN have higher triglycerides and cholesterol (moderate evidence) and have | **Yes** | **Yes** | **90% consensus to exclude** | **NO** | Lipids are tested and the QRISK score (10-year cardiovascular risk) is |
|  | cardiovascular risk | a 30% higher risk of CVD events (good evidence) compared to normotensive patients. The GP can manage this risk with lifestyle advice, tight blood pressure control with medication, and treatment with statins.<br>Conflicting evidence was found on the effect of lipid profile tests on behaviour (e.g. following lifestyle advice).<br>Regular lipid testing may be associated with a reduction in hospitalisations (weak evidence, target trial) |  |  |  |  | calculated at HTN diagnosis and the consensus group was not convinced that further monitoring is beneficial.<br>There is no good evidence to suggest that lipid monitoring promotes healthy life style changes. |
| **Liver function tests** | To detect metabolic dysfunction-associated steatotic liver disease (MASLD) | MASLD may be more common in people with HTN compared to the general population (weak conflicting evidence). Estimates vary (6-80% increased risk) and there is some contradictory evidence (no or negative association). | **No\*** | **Unclear** | 100% consensus to exclude this test. | **NO** | The consensus group decided that monitoring liver function does not benefit patient outcomes due to the lack of treatments for MASLD. |
| **Haemoglobin (full blood count)** | To detect anaemia | No evidence was identified on the risk of anaemia in patients with HTN. | **Yes** | **Unclear** | 88% consensus to exclude this test | **NO** | Anaemia can be treated, but it's unclear if intervention before symptom onset is beneficial. |
| **Thyroid function tests** | To detect hyper- or hypothyroidism | No evidence was identified on the risk of hyper- or hypothyroidism in patients with HTN. | Yes | Unclear | 86% consensus to exclude this test. | NO | The consensus group agreed that there is no rationale (known biological mechanism) to expect a difference in thyroid problems for hypertensive patients compared to the general population. |
| **Clotting tests** | To detect bleeding disorders | No evidence was identified on the risk of bleeding disorders in people with HTN. | Yes | Yes | 88% consensus to exclude this test. | NO | The consensus group agreed that there is no rationale (known biological mechanism) to expect a difference bleeding disorders for hypertensive patients compared to the general population. |
| **Adjusted calcium (bone profile)** | To detect hyper- or hypocalcaemia | No evidence was identified on the risk of abnormal (adjusted) calcium levels in people with HTN. | Yes | Unclear | 88% consensus to exclude this test. | NO | The consensus group agreed that there is no reason to suggest that patients with HTN are at a higher risk than the general population. |
| **Serum ferritin, folate acid and vitamin B12** | To detect deficiencies and anaemias | No evidence was identified on the risk of ferritin, folate, or vitamin B12 deficiencies in patients with HTN. | Yes | Unclear | 86% consensus to exclude this test. | NO | The consensus group decided that these tests are not required in the absence of evidence and a plausible biological mechanism. |
| **C-reactive protein (CRP) and erythrocyte sedimentation rate (ESR)** | General marker for inflammation (and infection), but are not linked to any particular condition | Yes (good evidence) | No (only lifestyle advice) | Unclear | 100% consensus to exclude | NO | There is no guidance on how to treat low grade inflammation. It is unclear whether reducing CRP or ESR directly has any benefit. |
| **Brain natriuretic peptide** | To detect heart failure | People with heart failure are more likely to have HTN compared to people without heart failure (weak evidence). |  |  | 88% consensus to exclude this test. | NO | The consensus group decided that the test has low accuracy for heart failure and was not necessarily developed as a monitoring test and should thus be excluded. |

\*This may change as new treatment become available for MASLD. Abbreviations: HTN: hypertension.

**Table 2.**
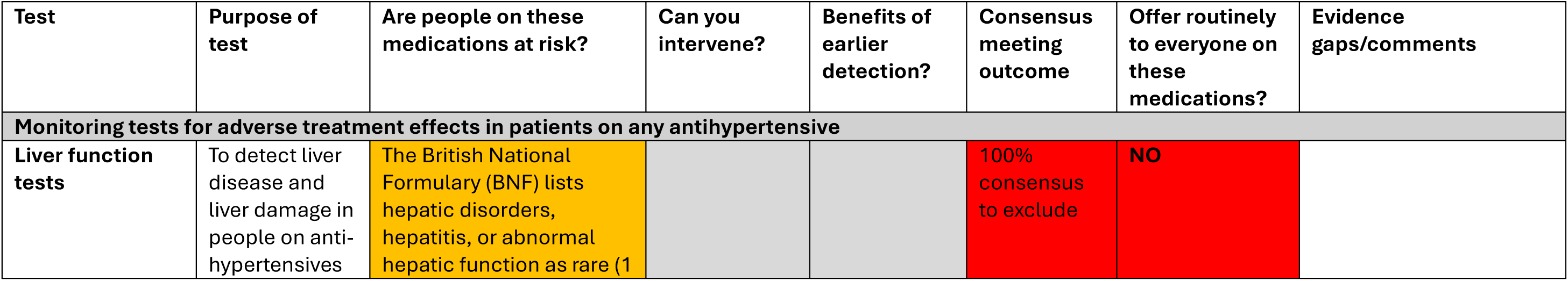
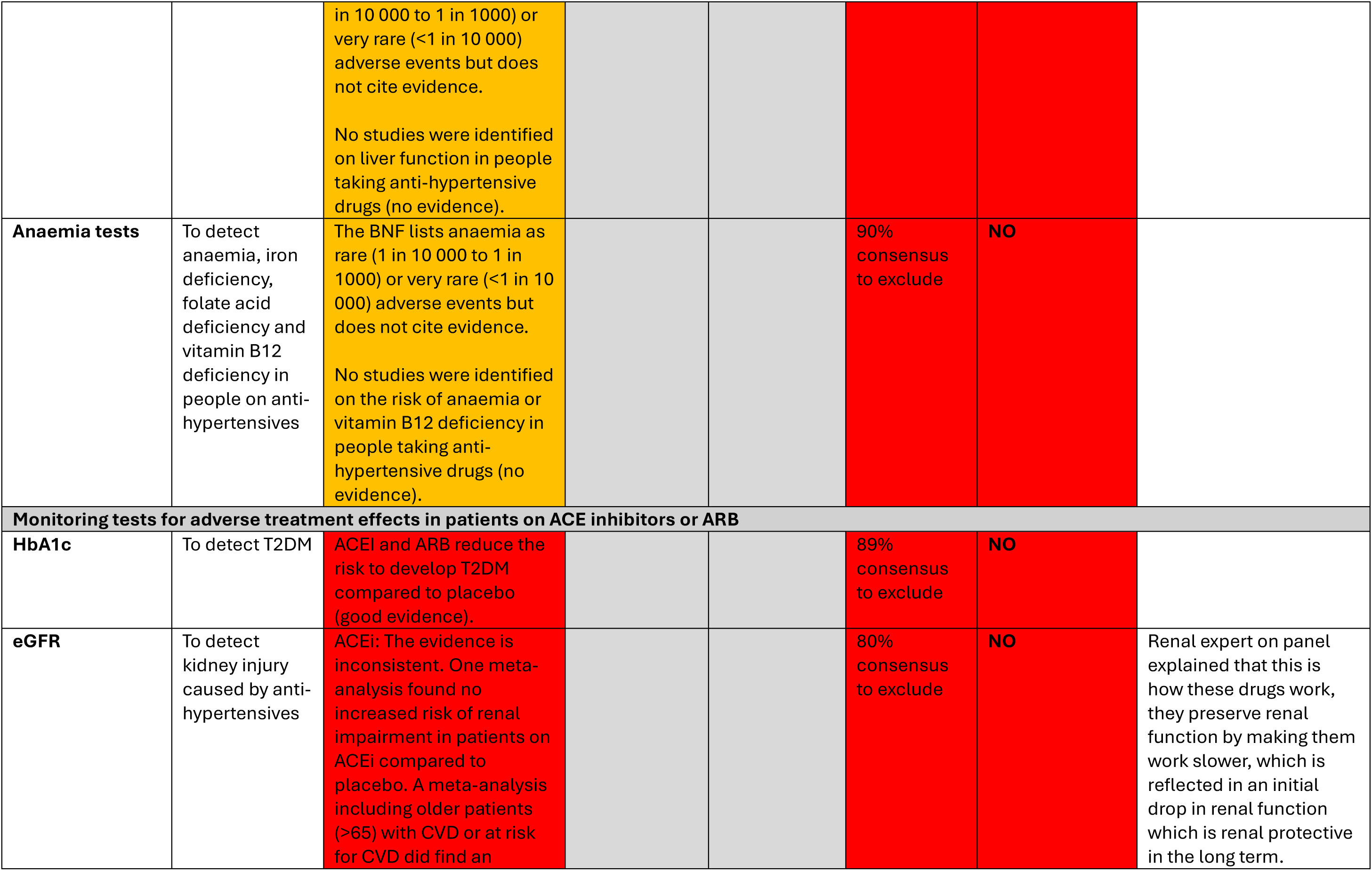
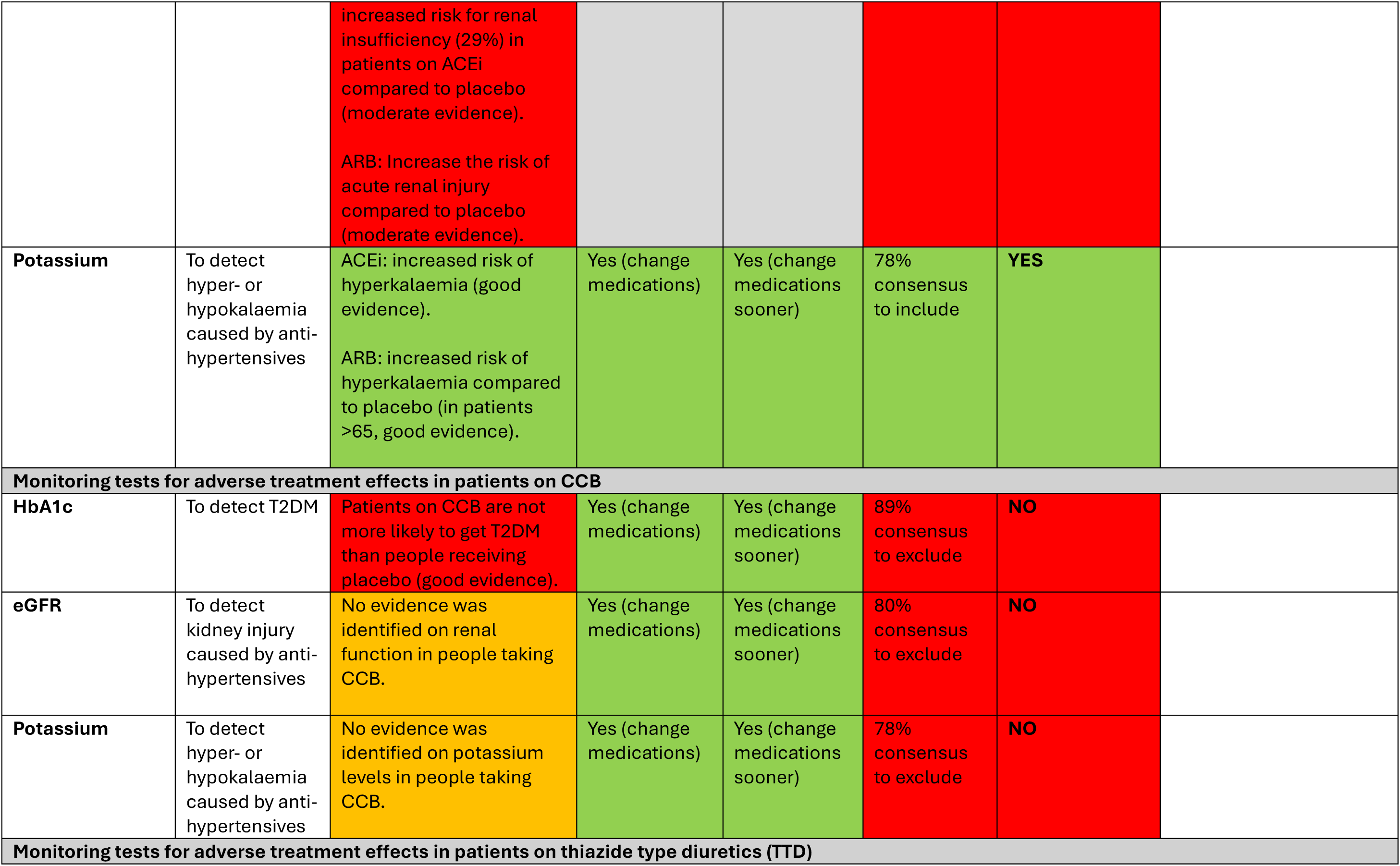
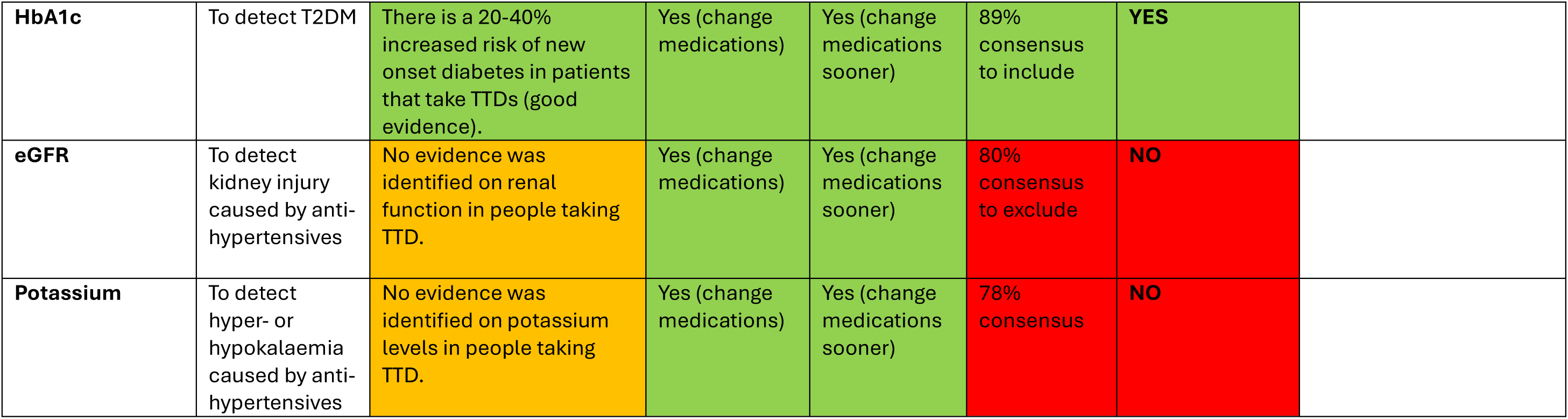
Tests to screen for adverse treatment effects of antihypertensives for patients on stable treatment – answers to filtering questions.

Five combined searches for eighteen different rapid reviews were conducted (see Supplementary Figure S1-5 for the PRISMA flow diagrams).

### Tests with good evidence and rationale

There is consistent, good evidence showing that people with hypertension are at higher risk of developing CKD than the general population.^16–20^ The consensus panel agreed that screening for CKD using eGFR is beneficial.

There is consistent, good evidence that people with hypertension are more likely to have T2DM, they are more likely to develop T2DM after diagnosis of hypertension, and increases and decreases in blood pressure are related to increased and decreased risk, respectively, of developing T2DM.^19–22^ Early detection and treatment of T2DM is beneficial,^23–27^ so the consensus panel agreed there is a strong rationale for regular monitoring of HbA1c.

Finally, we found good evidence that ACE inhibiters and ARBs increase the risk of hyperkalaemia,^28,29^ and for TTDs to increase the risk of T2DM,^30,31^ and there was consensus to include potassium and HbA1c for people on these medications, respectively.

### Tests with unclear evidence or rationale

We found inconsistent evidence whether hypertension increases the risk of hyper- or hypokalaemia. A systematic review including 17 studies found a higher prevalence of hyperkalaemia in hypertensive compared to normotensive people.^32^ Analyses of routine data showed that hypertensive patients with normal potassium levels at diagnosis have a similar incidence of abnormal levels compared to controls.^15^

We found no evidence on the prevalence of abnormal serum sodium levels in hypertensive patients. Analyses of routine data showed that hypertensive patients who had normal sodium levels at diagnosis were at higher risk to develop abnormal levels compared to controls.^15^ However, this may have been caused by medications rather than hypertension itself, which was not taken into account in this analysis. Thus, this did not provide strong evidence that people with hypertension who do not take medication benefit from sodium monitoring.

Based on this limited evidence there was consensus to exclude both potassium and sodium from the final panel, at least for people not on any medications. However, this is unlikely to change clinical practice, since most labs will automatically measure sodium and potassium if eGFR is requested. In addition, many patients will be on medications, which may change the decision to monitoring potassium and sodium.

Lipid profile is used to calculate the Q-RISK score to assess cardiovascular risk and there is good and consistent evidence from multiple systematic reviews that hypertension increases the risk of cardiovascular disease (CVD).^33–35^ In addition, a large population based survey has shown that hypertension is associated with higher lipid levels. However, the Q-RISK score can also be calculated without lipids or with a single measurement of lipids at diagnosis. The GP can manage this risk with lifestyle advice, tight blood pressure control with medication, and treatment with statins. The consensus group suggested that regular lipid monitoring may motivate people to adhere to life style advice, but the evidence on this is inconsistent and weak, suggesting that this is not a good rationale for monitoring. Given the available evidence, the consensus group was not convinced that regular monitoring after initial assessment of lipid profile at diagnosis is beneficial and excluded the test from the panel.

No evidence was identified whether hypertension increases the risk of conditions that can be picked up by haemoglobin, thyroid function, clotting, calcium, ferritin, folate acid, and vitamin B12 tests. The consensus group decided that in the absence of evidence and a known biological mechanism, these tests should not be included.

No evidence was identified linking antihypertensive drugs to abnormal liver function or anaemia once patients are on stable treatment. Also no evidence was identified on eGFR or potassium levels in people taking CCB or TTD. In the absence of evidence of clinical need, the consensus group decided to exclude these tests for the purpose of monitoring adverse effects of these medications.

### Tests that should not be routinely offered to people with hypertension

Liver function tests can help to detect metabolic dysfunction-associated steatotic liver disease (MASLD), but it is unclear if people with hypertension are at greater risk because the evidence is weak and contradictory.^36–38^ Importantly, the management, namely lifestyle advice, would be similar to what the patients already receive for hypertension management. There was consensus that monitoring liver function does not benefit patient outcomes due to the lack of treatments for MASLD, although it is important to note that this may change in the future.

CRP and ESR were excluded because there is no guidance or agreement on how to treat low grade inflammation. Brain natriuretic peptide was excluded because it was not developed as a monitoring test and was believed to be too inaccurate.

We found good evidence that ACE inhibiters and ARBs are renoprotective and reduce the risk of developing T2DM,^30,31^ so additional monitoring of eGFR and HbA1c in people on these medications is not warranted. We also found good evidence that people on CCB are not more likely to get T2DM than people receiving placebo,^30,31^ so additional monitoring HbA1c in people on CCB is not warranted either.

## DISCUSSION

### Summary

We identified a minimal evidence-based testing panel of blood tests for adults with hypertension. This panel includes eGFR to screen for CKD, HbA1c to screen for T2DM, potassium for patients on ACE inhibiters and ARBs. We also identified a list of tests where evidence is absent or inconsistent: lipids, electrolytes, haemoglobin, thyroid function tests, clotting tests, calcium, ferritin, folate acid, or vitamin B12 to screen for secondary conditions; liver function and anaemia tests for adverse effects caused by antihypertensives, and eGFR or potassium levels for adverse effects caused by CCB or TTD. Finally, we found good evidence that there is no benefit in routinely monitoring liver function, inflammation markers such as CRP and ESR, or brain natriuretic peptide and that monitoring eGFR and HbA1c to detect side effects of ACE inhibiters, ARBs, and CCB is not warranted. The consensus group agreed that in the absence of evidence or clinical need these tests should not be offered to people with hypertension.

### Strengths and limitations

The main strength of this study is that we used established and systematic methods to identify evidence. To address some of the gaps in the literature, we conducted routine data analyses using a large cohort of primary care patients who are largely representative of the UK population regarding age, ethnicity, and sex. Another strength is that we included a diverse consensus group with primary care nurses, GPs, consultants, and patient representatives to judge the evidence and make the final decisions on including or excluding tests for the minimal testing panel.

This study also has several limitations. For pragmatic reasons, we used stepwise rapid review methods, so as a result we may have missed relevant evidence. However, we used standard rapid review methods and we only stopped the search once we found good quality evidence. For most tests no (good) evidence was found, so all searches were performed and all records were screened. We did not consider the accuracy, biological variation, or optimal thresholds of each tests when considering the evidence, which may be important to consider. However, the few tests that were included in the final panel are well established. Although we included diverse members in the consensus group, their opinion may not reflect the wider community. We also did not include a health economic evaluation in this analysis, and we have not looked at the optimal frequency of monitoring which are important next steps to address.

### Comparison with existing literature

Similar to current UK guidelines, who recommend regular renal function testing, we recommend regular testing of eGFR, although we do not recommend regular testing of electrolytes.^39^ However, many laboratories will include electrolytes automatically when ordering eGFR, so this recommendation may have limited impact. We recommend regular monitoring of HbA1c to detect new onset T2DM in people with hypertension. This is not mentioned in NICE Hypertension guidelines,^39^ although it is suggested in the published health diabetes prevention guidelines.{NICE, 2017 #116}

In practice, the majority of patients receive more tests than are in our minimal testing set or that are recommended by guidelines. Many patients with hypertension receive annual monitoring of full blood count, lipid profile, and liver function.^7^ A survey amongst GPs suggest that this is not just due to clinical need or comorbidities, because it showed that 19-52% of GPs would routinely order lipids, liver function tests, and full blood count for an average hypertensive patient.^8^

### Implications for research and practice

Our study looked for evidence supporting the rationale of the use of a test, but did not consider optimal frequency of monitoring. Many tests are offered annually for practical reasons but it is unclear if this is the best frequency. We also did not consider optimal thresholds, which may be different from how tests are currently used. Future research is needed to address these gaps. As randomised trials on monitoring are challenging, due to the long follow-up time that is needed to show the benefit of monitoring, these gaps may be partially addressed using health economic modelling.

The evidence-base for the majority of tests that are routinely ordered for adults with hypertension is weak or absent. In contrast, there is good evidence of the harms of over-testing, so in the absence of evidence, we suggest tests should not be offered routinely. National guidelines should reflect this and local protocols need to be updated to avoid unnecessary testing. New guidance and local protocols need to make clear that additional tests can be added if clinically indicated, but should not be included for routine monitoring purposes. Educating GPs and staff requesting tests about the harms of over-testing may help avoid tests being added “just in case” or “because we have always done this”.

Our research suggests that the minimal testing panel is beneficial for most hypertensive patients. However, there may be exceptions. In some patients, the benefit of monitoring may not outweigh the harms of testing. For instance, in frail elderly patients, who have multiple conditions and limited life expectancy, regular monitoring may do more harm than good.

Standardising testing for hypertension can address unwarranted variation in care, but the final decision to monitor, what to monitor and how often should still be made on individual basis and in collaboration with the patient.

## Conclusion

We identified a minimal set of evidence-based tests that should be offered to adults with hypertension. Implementing these recommendations could reduce harms associated with unwarranted variation in care. Further research is needed to clarify the role of tests with inconsistent evidence and determine the optimal frequency of testing.

## Supporting information

Supplementary materials - Evidence report

PRISMA checklist

## Data Availability

All data produced during the rapid reviews and consensus meetings are contained in the manuscript or supplementary materials.
Part of the study is based on data from the Clinical Practice Research Datalink (CPRD) obtained under licence from the MHRA and NIHR. The data are provided by patients and collected by the NHS as part of their care and support. CPRD data can be accessed via the MHRA: https://www.cprd.com/

## ACKNOWLEDGMENTS

We would like to express our gratitude to Aisling O’Rourke for her invaluable administrative support throughout the project and for organizing the consensus meeting. We also extend our sincere thanks to the members of the consensus group, including patient representatives Bob Cottis, Jean Palmer, and Patricia MacCalla; GPs Katherine Alsop, Sam Merriel, Rachel Johnson, and Alan McFarlane; renal consultant Dominic Taylor; and primary care nurse Jennifer Charlewood.

Second consensus meeting: patient representatives Bob Cottis, Jean Palmer, and Patricia MacCalla; GPs David Spitzer, Nazmul Mohsin, Tim Johnson, Sam Merriel, and Alan McFarlane; renal consultant Dominic Taylor; and primary care nurses Jennifer Charlewood and Helen Lane.

Additionally, we wish to acknowledge the contributions of the wider team, including Mary Ward, Howard Thom, Alice Malpass, Jonathan Banks, Clare Thomas, Hayley Jones, and Jonathan Sterne, as well as patient representatives Francesco Palma and Christina Stokes.

## AUTHORSHIP AND CONTRIBUTIONS

Martha MC Elwenspoek drafted the manuscript and developed the protocol. Rachel O’Donnell and Catalina Lopez Manzano conducted the rapid reviews, including abstract screening, full-text screening, data extraction, risk of bias assessment, and wrote the evidence report. Sarah Dawson was responsible for developing all search strategies, Lewis Buss conducted CPRD analyses, and Katie Charlwood identified evidence gaps. Alastair Hay and Jessica Watson contributed clinical expertise and secured funding for the project. Penny Whiting developed the methods, secured funding, and together with Jessice Watson provided project oversight.

All authors had full access to the data in the study and can take responsibility for the integrity and accuracy of the data analysis. All authors have read and approved the final manuscript.

## DATA SHARING

The data that support the findings of this study are available within this paper and supplementary files.

## ETHICS APPROVAL

Not applicable.

## CONSENT TO PARTICIPATE

Not applicable.

## CONSENT FOR PUBLICATION (FROM PATIENTS/PARTICIPANTS)

Not applicable.

## CODE AVAILABILITY

Not applicable.

