## Supplementary materials - Evidence report for "Evidence-based blood tests for monitoring adults with hypertension in primary care: rapid review, routine data analyses, and consensus study"

### Evidence report: Hypertension

#### Consensus meeting 2

Version 4

#### Report Summary

##### Purpose of the consensus meeting

We selected 11 test panels to consider for a routine tests for people with Hypertension (HTN). The purpose of the consensus meeting is to decide which tests should be included for a **minimal testing panel** for people with HTN, which tests can be removed from this panel, and for which tests more evidence is needed before a decision can be made.

This minimal testing panel is aimed at an adult that has been diagnosed with a long term condition without complications. This panel lists tests that should be used to regularly monitor this long term condition as standard.

##### Changes made due to the previous consensus feedback

Tests for both C-reactive protein (CRP) and erythrocyte sedimentation rate (ESR) are general markers for inflammation (and infection). It was decided at the previous consensus meeting where T2DM was discussed, that these tests should be removed from future panels as there is no guidance on how to treat low grade inflammation and the tests do not point to a reason for inflammation.

For this report, each testing panel has been split into its constituent individual tests where possible; an overall evidence rating has been given to each individual test and to the testing panel as a whole. We will initially vote on whether to include the "testing panel"; where panels are voted for inclusion or for further evidence, we will then vote on each individual test.

##### Method for Evidence Rapid Reviews

1. Tests were selected based on:
    a. Tests that are currently ordered for patients with Hypertension (HTN) in primary care (CPRD analysis)
    b. Tests that GPs say they would order for the average HTN patient (online survey of GPs).
    c. Tests that are recommended by UK guidelines for routine monitoring of patients with HTN.
2. Tests were categorised as
    a. Tests to monitor disease progression and treatment response
    b. Tests to screen for secondary conditions
    c. Tests to screen for adverse treatment effects

3. We determined a list of filtering questions that needed to be answered with ‘yes’ for a test to be a useful monitoring test.
    a. For secondary condition monitoring:
        I. Which acute or chronic complications do these tests pick up?
        II. Is X (or abnormal X tests) more common in LTC patients compared to the general population?
        III. Is there anything the GP can do to manage or treat X?
        IV. Are there clear benefits of earlier detection or treatment?
    b. For drug side effect monitoring:
        I. Is X more common in people who take antihypertensive drugs?
        II. Is there anything the GP can do to manage or treat X?
        III. Are there clear benefits of earlier detection or treatment?
4. We looked at the following sources and extracted any evidence in favour or against using each test. We only proceeded to the next source if insufficient evidence was found.
    a. NICE guidelines and NICE clinical knowledge summaries (including references) (CKS)
    b. Systematic reviews
    c. Primary studies in large primary care datasets, cohorts, or relevant trials.

#### Long Term Condition Monitoring Evidence Summary

There are no blood tests related to the monitoring of hypertension itself. Monitoring involves blood pressure checks, which is beyond the scope of our project which focuses only on routine blood tests.

#### Tests to Screen Secondary Conditions Summary

11 testing panels were included to screen for secondary conditions related to HTN. These testing panels were broken down into singular tests within the panel and given and overall evidence rating which will be presented at the consensus meeting for consideration. Evidence was gathered for both prevalence of condition and evidence of using singular tests to test for secondary condition. (see table 1 for summary)

#### Tests to monitor adverse treatment effects

There are many anti-hypertensive drugs on the market, but the first line treatments are angiotensin-converting enzyme (ACE) inhibitors, Angiotensin II receptor blockers (ARB), thiazide type diuretic (TTD) and calcium channel blockers (CCB). Prescribing additional drugs is often used when patients have resistant hypertension which makes up less than 20% of patients and often confounded by non-adherence. Therefore, we only considered side effects of these 4 groups of drugs, and we only considered side effects that are listed on British National Formulary (BNF). (see table 2 for summary)

#### Evidence Rating Guide

We labelled evidence found in the rapid reviews as follows:

| **Rapid review evidence rating** | |
|---|---|
| Good evidence | High quality systematic reviews or high quality studies with large sample sizes |
| Moderate evidence | A systematic review with some quality concerns or several primary studies showing similar results but some studies have quality concerns. |
| Weak evidence | A single primary study with a small sample size with or without quality concerns. |
| Conflicting evidence | Evidence found has contradicting findings. |
| No evidence found | No evidence was found for the condition, adverse effect or single test during the rapid reviews. |
| Unsure | Further discussion is needed in the consensus meeting about the test. |

#### Table 1: Summary Evidence for tests to screen for secondary conditions

*Please click the Blood tests panel names to be taken to the corresponding evidence further down this report.*

| Blood Test Panel | Related secondary condition | Evidence rating for condition | Individual test types within testing panels | Evidence rating for single test | Treatable/ manageable by GP? | Benefit to earlier detection? |
|---|---|---|---|---|---|---|
| **HbA1C** | Type 2 diabetes | Good | HbA1C | Good | Yes | Yes |
| **Renal Function tests** | Chronic kidney disease | Good | eGFR | Good | Yes | Yes |
| | | | Blood urea | No evidence | Yes | Yes |
| **Blood electrolyte tests** | Hyper- or hypokalaemia | Moderate | Serum potassium | Moderate | Yes | Yes |
| | Hyper- or hyponatremia | No evidence | Serum sodium | No evidence | No | Yes |
| **Liver function tests** | Non-alcoholic fatty liver disease | Weak (conflicting evidence) | Albumin | No evidence | No | No |
| | | | Alkaline phosphatase (ALP) | No evidence | No | No |
| | | | Alanine aminotransferase (ALT) | No evidence | No | No |
| | | | Aspartate aminotransferase (AST) | No evidence | No | No |
| | | | Bilirubin | No evidence | No | No |
| | | | Gamma glutamyl transpeptidase (GGT) | Good | No | No |
| | | | Total protein | No evidence | No | No |
| **Lipid Profile** | Dyslipidaemia, cardiovascular disease risk | Weak for Dyslipidaemia, | Serum cholesterol | Good | Yes | Yes |
| | | | High-density lipoprotein (HDL) | Good | Yes | Yes |
| | | Good for CVD Risk | Low-density lipoprotein (LDL) | Unsure | Yes | Yes |
| | | | HDL:LDL ratio | Unsure | Yes | Yes |
| | | | Triglycerides | Weak | Yes | Yes |
| **Full blood count** | Anaemia | No evidence | Haemoglobin (Hb) | No evidence | Yes | Yes |
| **Haematinics** | Anaemia and deficiency type | No evidence | Vitamin B12 | No evidence | Yes | Yes |
| | | | Serum ferritin | No evidence | Yes | Yes |
| | | | Folate tests | No evidence | Yes | Yes |
| **Thyroid function Tests** | Hyper- or hypothyroidism | No evidence | Thyroxine (T4) | No evidence | Yes | Yes |
| | | | Free thyroxine (Free T4) | No evidence | Yes | Yes |
| | | | Thyroid stimulating hormone | No evidence | Yes | Yes |
| | | | Free triiodothyronine (T3) | No evidence | Yes | Yes |
| | | | Serum T3 level | No evidence | Yes | Yes |
| **Clotting Tests** | Bleeding disorders | No evidence | Prothrombin time | No evidence | No evidence | No evidence |
| | | | Partial thromboplastin time | No evidence | No evidence | No evidence |
| | | | Thrombin time | No evidence | No evidence | No evidence |

| Blood Test Panel | Related secondary condition | Evidence rating for condition | Individual test types within testing panels | Evidence rating for single test | Treatable/ manageable by GP? | Benefit to earlier detection? |
|---|---|---|---|---|---|---|
| **Bone Profile** | Late-stage kidney disease, bone disorders, serum phosphate levels, and parathyroid disorders | Weak evidence found for vitamin D deficiency | Serum inorganic phosphate | No evidence | Yes | No evidence |
| | | | Alkaline phosphatase | No evidence | Yes | No evidence |
| | | | Calcium | No evidence | Yes | No evidence |
| | | | Calcium (adjusted) | No evidence | Yes | No evidence |
| | | | Parathyroid hormone | No evidence | Yes | No evidence |
| | | | Vitamin D | Weak | Yes | No evidence |
| **B-type natriuretic peptide (BNP)** | Heart failure | Weak | BNP | Weak | Yes | No evidence |

#### Table 2: Summary Evidence for Drug Adverse Effect Monitoring

*Please click the Blood tests panel names to be taken to the corresponding evidence further down this report.*

| Blood Tests Panel | Adverse effect (AE) | Evidence rating for AE | Individual test types within testing panels | Evidence rating for single test and adverse effect | Treatable/ manageable by GP? | Benefit to earlier detection? |
|---|---|---|---|---|---|---|
| **Blood glucose** | Increased blood glucose or new onset diabetes | Moderate | HbA1c | No evidence | Yes | Yes |
| **Renal function tests** | Kidney injury | Moderate (some conflicting evidence found) | eGFR (serum creatine) | Weak | No | Yes |
| | | | Blood urea | No evidence | No | Yes |
| | | | Serum potassium | Moderate | No | Yes |
| | | | Serum Sodium | No evidence | No | Yes |
| **Liver function tests** | Liver injury | No evidence | Albumin | No evidence | No | Yes |
| | | | Alkaline phosphatase (ALP) | No evidence | No | Yes |
| | | | Alanine aminotransferase (ALT) | No evidence | No | Yes |
| | | | Aspartate aminotransferase (AST) | No evidence | No | Yes |
| | | | Bilirubin | No evidence | No | Yes |
| | | | Gamma glutamyl transpeptidase (GGT) | No evidence | No | Yes |
| | | | Total protein | No evidence | No | Yes |
| **Anaemia** | Anaemia | No evidence | Haemoglobin (Hb) | No evidence | Yes | Yes |
| | | | Vitamin B12 | No evidence | Yes | Yes |
| | | | Serum ferritin | No evidence | Yes | Yes |
| | | | Folate tests | No evidence | Yes | Yes |

### Tests to screen for secondary conditions

#### 1. HbA1C

##### 1.1 Which acute or chronic complications do these tests pick up?

Type 2 diabetes mellitus (T2DM) (NICE guidance)

##### 1.2 Is T2DM (or abnormal blood glucose tests) more common in HTN patients compared to the general population?

Yes, type 2 diabetes is more common in hypertensive patients compared to normotensive patients, with one study suggesting that a 20 mm/Hg increase from normal systolic blood pressure increasing diabetes prevalence.

| Source | Blood Tests | Evidence | Quality concerns |
|---|---|---|---|
| NICE Guidelines | HbA1C | CKS recommends for investigations into target organ damage for hypertension, this includes arranging measurement of HbA1C (to test for diabetes).<br>https://cks.nice.org.uk/topics/hypertension/diagnosis/investigations/ | |
| *Systematic reviews* | | | |
| Emdin 2015 (1) | No specific tests mentioned for diabetes | • Population: Global and European populations<br>• Sample size: 30 prospective observational studies (n = 285,664 with 17,388 incident diabetes events).<br>• Findings:<br>○ Pooled random effects meta-analysis showed that with a 20 mm/Hg increase from usual systolic blood pressure (SBP) was associated with a 77% higher risk of new diabetes. Relative risk = 1.77 [95% CI 1.53- 2.05].<br>**Conclusion: Higher levels of SBP increase the risk of developing diabetes, with a 20 mm/Hg increase from the normal SBP increasing the risk by 77%** | Limited search strategy. Risk of bias of individual studies was not analysed. |
| *Primary Studies* | | | |
| Muntner 2022 (2) | No specific tests mentioned for diabetes | • Population: NHANES (2011-2018), participants aged >18.<br>• Sample size: 21,518 (hypertension: n = 14,158, non-hypertension: n = 7,360).<br>• Findings:<br>○ Prevalence of diabetes in patients aged 18-44: hypertensive: 5.7% (n = 3,962), normotensive: 1.3% (n = 5,549).<br>○ Prevalence of diabetes in patients aged 45-64: hypertensive: 18.8% (n = 5,624), normotensive: 4.2% (n = 1,565).<br>○ Prevalence of diabetes in patients aged 65+: hypertensive: 24.5% (n = 4,572), normotensive: 7% (n = 309).<br>**Conclusion: Diabetes is more common in hypertensive patients than in normotensive patients, across all age groups.** | Self-reported diabetes |
| Hua 2021 (3) | No specific tests | • Population: NHANES (2007-2018) participants aged 18-80.<br>• Sample size: 26,778, participants (hypertensive (n = 13,283), normotensive: (n = 13,495) | Self-reported diabetes. |

| Source | Blood Tests | Evidence | Quality concerns |
|---|---|---|---|
|  | mentioned for diabetes | • Findings:<br> ○ Prevalence of diabetes: hypertensive: 18.8%, normotensive: 4.9%<br>**Conclusion: Diabetes is more common in hypertensive patients than in normotensive patients.** |  |
| Emdin 2015 (1) | No specific tests mentioned for diabetes | • Population: CPRD patients aged 30-90.<br>• Sample size: 4,132,138 (SBP groupings: < 127 mm/Hg (n = 1,474,462), 127-136 mm/Hg (n = 1,541,802), and > 136 mm/Hg (n = 1,115,874)).<br>• Findings:<br> ○ A 20 mm/Hg increase in SBP, and a 10 mm/Hg increase in DBP were associated with a 58% increase (hazard ratio (HR) = 1.58[95% CI 1.56-1.59] and 52% increase (HR = 1.52[95% CI 1.51-1.54] in risk of newly diagnosed diabetes, respectively.<br>**Conclusion: 20 mm/Hg increase in SBP increases the risk of diabetes by 58%, 10 mm/Hg increase in DBP increase the risk of diabetes by 52%.** | No |

#### 1.3 Is there anything the GP can do to manage or treat T2DM?

Yes, lifestyle changes and drug treatment to control blood glucose levels. There are several options for medications which can be changed in dose or combination. Answered in the type 2 diabetes mellitus evidence report and presented in the Feb 2022 consensus meeting.

#### 1.4 Are there clear benefits of earlier detection or treatment?

Yes, early intervention with drugs has long term benefits. Answered in the type 2 diabetes mellitus evidence report and presented in the Feb 2022 consensus meeting.

#### 2. Renal function tests (eGFR and blood urea)

##### 2.1 Which acute or chronic complications do these tests pick up?

Chronic kidney disease (CKD) (NICE guidance, ICHOM)

##### 2.2 Is CKD (or abnormal renal function tests) more common in HTN patients compared to the general population?

Yes, the risk of kidney disease is 2 to 3 fold higher in those with hypertension compared to those without.

| Source | Blood Tests | Evidence | Quality concerns |
|---|---|---|---|
| NICE Guidelines | eGFR | Hypertension increases the risk of several conditions, including CKD.<br>https://cks.nice.org.uk/topics/hypertension/background-information/complications-prognosis/<br>At annual review: Check renal function by measuring serum creatinine, electrolytes, and estimated glomerular filtration rate (eGFR), and urine to check for albumin: creatinine ratio (ACR).<br>https://cks.nice.org.uk/topics/hypertension/management/management/ | |
| *Systematic reviews* | | | |
| Shrestha 2021 (4) | eGFR | • Population: South Asian<br>• Sample size: general population (n = 50,494, 16 studies), hypertensive adults (n = 22,973, 12 studies).<br>• Findings:<br>o The pooled prevalence of CKD among the general population was 14% [95% CI 11–18%],<br>o The prevalence of CKD was 27% [95% CI 20–35%] in hypertensive adults.<br>**Conclusion: The prevalence of CKD is nearly double in those with hypertension compared to the general population.** | Inadequate search, high heterogeneity without subgroup analysis, some evidence of publication bias, and only south Asian population was included in this study. |
| Zhang 2019 (5) | No specific tests mentioned for renal disease | • Population: patients with proven masked hypertension and normotensive individuals.<br>• Sample size: 21 studies (n = 130,318), normotensive individuals (n = 15,872), controlled hypertension (n = 21,176), masked hypertension (n = 5,706), and masked uncontrolled hypertension (n = 12,462).<br>• Findings:<br>o The pooled risk ratio for masked hypertension vs normotension was 3.85 [95% CI 2.03–7.31] for renal disease events<br>**Conclusion: The risk of CKD is over 3-fold for those with hypertension compared to normotensive individuals.** | No |
| Garofalo 2016 (6) | eGFR | • Population: adults with prehypertension, hypertension or normotension.<br>• Sample size: 16 studies included, normotensive individuals (n = 15,872), controlled hypertension (n = 21,176), masked hypertension (n = 5,706), and masked uncontrolled hypertension (n = 12,462).<br>• Findings:<br>o Presence of prehypertension and hypertension increased renal risk (RRs of 1.19 [95% CI 1.07-1.33] and 1.76 [95% CI 1.58-1.97], respectively). | No |

| Source | Blood Tests | Evidence | Quality concerns |
|---|---|---|---|
| | | ○ Every 10 mm/Hg increase in systolic and diastolic blood pressure was associated with higher risk for decreased eGFR (RRs of 1.08 [95% CI 1.04-1.11] and 1.12 [95% CI 1.04-1.20], respectively).<br>**Conclusion: The risk of CKD is nearly 2 fold for those with hypertension.** | |
| *Primary studies* | | | |
| Muntner 2022 (2) | eGFR | • Population: NHANES (2011-2018), participants aged >18.<br>• Sample size: 21,518 total (hypertension: n = 14,158, non-hypertension: n = 7,360).<br>• Findings:<br>○ Prevalence of reduced eGFR in patients aged 18-44: hypertensive: 0.8% (n = 3,962), normotensive 0.1% (n = 5,549).<br>○ Prevalence of reduced eGFR in patients aged 45-64: hypertensive: 5.1% (n = 5,624), normotensive: 2.5% (n = 1,565).<br>○ Prevalence of reduced eGFR in patients aged 65+: hypertensive: 26.8% (n = 4,572), normotensive: 15% (n = 309).<br>**Conclusion: Reduced eGFR is more common in hypertensive patients than in normotensive ones, across all age groups.** | No |
| Hua 2021 (3) | eGFR | • Population: NHANES (2007- 2018), participants aged 18-80.<br>• Sample size: 26,778, 49.6% of patients had hypertension.<br>• Findings:<br>○ eGFR (ml/min/1.73m$^2$)<br>▪ Non=hypertensive: 118.7 (n = 13,495).<br>▪ Hypertensive; 109.2 (n = 13,283).<br>**Conclusion: eGFR is lower in hypertensive patients compared to normotensive patients.** | No |

#### 2.2 Is there anything the GP can do to manage or treat CKD?

Answered in the CKD evidence report that will be presented alongside this report in the Feb 2023 consensus meeting.

#### 2.3 Are there clear benefits of earlier detection or treatment?

Answered in the CKD evidence report that will be presented alongside this report in the Feb 2023 consensus meeting.

#### 3. Blood Electrolyte tests (Serum potassium and serum sodium)

##### 3.1 Which acute or chronic complications do these tests pick up?

Hyper- or Hypokalaemia and Hyper- or Hyponatremia. (CKS)

##### 3.2 Are abnormal blood electrolyte levels more common in HTN patients compared to the general population?

The prevalence of serum potassium exceeding 5.0 mmol/L is higher in hypertensive patients than in normotensive patients. No evidence was found for the prevalence of abnormal serum sodium levels in hypertensive patents compared to the general population.

| Source | Blood Tests | Evidence | Quality concerns |
|---|---|---|---|
| NICE Guidelines | Electrolytes (potassium and sodium) | At annual review: Check renal function by measuring serum creatinine, electrolytes, and estimated glomerular filtration rate (eGFR), and urine to check for albumin: creatinine ratio (ACR). https://cks.nice.org.uk/topics/hypertension/management/management/ | |
| *Systematic reviews* | | | |
| Palaka 2020 (7) | Serum potassium | • Population: Adult patients with CKD, heart failure, type 2 diabetes, or hypertension.<br>• Sample size: 123 studies included, with 17 studies on hypertension.<br>• Findings:<br> ○ Studies with cohorts that included only Hypertensive patients reported prevalence of serum potassium concentrations exceeding 5.0 mmol/L ratios between 1.06 and 1.14.<br>**Conclusion: The prevalence of serum potassium exceeding 5.0 mmol/L is higher in hypertensive patients than in normotensive patients.** | No mention of a risk of bias assessment for the included studies. Data was not pooled, and no indication of drugs taken into account. |
| *Primary studies* | | | |
| None identified | | | |

#### 3.3 Is there anything the GP can do to manage or treat abnormal blood electrolyte levels?

Yes, mild or moderate hyperkalaemia may be managed by dietary modification or medication changes for those with hypertension. However, in the event of life-threatening acute hyperkalaemia, emergency hospital admission may be needed, and drugs are available to reduce potassium levels to a safer range.

| Source | Evidence |
|---|---|
| NICE | Sodium zirconium cyclosilicate for treating hyperkalaemia<br>NICE Technology appraisal guidance [TA599]<br>Published: 24 January 2022<br>https://www.nice.org.uk/guidance/ta599/resources/sodium-zirconium-cyclosilicate-for-treating-hyperkalaemia-pdf-82607272135621<br><br>Patiromer for treating hyperkalaemia<br>NICE Technology appraisal guidance [TA623]<br>Published: 13 February 2020<br>https://www.nice.org.uk/guidance/ta623/resources/patiromer-for-treating-hyperkalaemia-pdf-82609015577029<br>Both State:<br>**Treatment for hyperkalaemia depends on its severity. Life-threatening acute hyperkalaemia needs emergency treatment in hospital.**<br>Small rises in serum potassium above this can cause electrocardiogram (ECG) changes. To lower the risk of cardiac arrest, clinicians use active potassium-lowering treatments, then identify and remove the cause of hyperkalaemia. The guidelines include the following treatments:<br>• calcium chloride or calcium gluconate intravenously to protect the heart if there is ECG evidence of hyperkalaemia<br>• insulin and glucose intravenously to move potassium from the blood into cells<br>• nebulized salbutamol as an adjunctive therapy to insulin and glucose for serum potassium levels of 6.5 mmol/litre and above to move potassium from the blood in to cells<br>• after severe hyperkalaemia has resolved, potassium-binding agents for 3 or more days (namely, calcium resonium given orally) to remove potassium from the body<br>• stopping or reducing RAAS inhibitors, which can increase serum potassium levels.<br><br>**The aim of treatment for chronic hyperkalaemia is to lower potassium levels to prevent acute life-threatening hyperkalaemia. Treatment includes:**<br>• advising people with chronic kidney disease to avoid foods high in potassium<br>• stopping or reducing RAAS inhibitors and potassium-sparing diuretics<br>• avoiding non-steroidal anti-inflammatory drugs and trimethoprim. |

#### 3.4 Are there clear benefits of earlier detection or treatment?

Yes. Late detection of high potassium levels can lead to an increased risk of cardiac arrest.

#### 4. Liver function tests (albumin, alkaline phosphatase (ALP), alanine aminotransferase (ALT), aspartate aminotransferase (AST), bilirubin, gamma glutamyl transpeptidase (GGT), total protein)

##### 4.1 Which acute or chronic complications do these tests pick up?

Non-alcoholic fatty liver disease (NAFLD) (including non-alcoholic fatty liver (NAFL) and non-alcoholic steatohepatitis (NASH)) (NICE)

##### 4.2 Is NAFLD (or abnormal liver test results) more common in HTN patients compared to the general population?

Yes, NAFLD is nearly twice as likely in hypertension compared to the general population, there is some evidence to the contrary.

| **Source** | **Blood Tests** | **Evidence** | **Quality concerns** |
|---|---|---|---|
| NICE Guidelines | | Not reported | |
| *Systematic Reviews* | | | |
| Jarvis 2020 (8) | Abnormal liver blood tests, no specific tests mentioned. | • Population: Adults with and without metabolic risk factors.<br>• Sample size: 22 studies (16 cohorts, 4 of these studies reported on hypertension)<br>• Findings:<br>○ 1 study found a negative association after adjustment for other metabolic risk factors between hypertension and NAFLD (HR 0.07 [95% CI 0.01–0.3]).<br>○ This is contradicted by 2 larger population-based data linkage studies that both report a positive association between diagnosed hypertension and an incident liver outcome (HR: 1.23 [95% CI 1.14–1.31] and 1.59 [95% CI 1.51–1.69]).<br>○ The final study, which used several large European primary care datasets, reported a smaller positive association between hypertension and non-fatal liver outcomes (HR 1.06 [95% CI 1.00–1.12; p = 0.03]).<br>**Conclusion: The prevalence of NAFLD is uncertain and contradicted in this review, but the suggestion from the largest, highest quality studies was that hypertension is associated with incident severe liver disease.** | Authors did not believe it was appropriate to pool data, so no meta-analysis was run. |
| *Primary Studies* | | | |
| Younossi 2020 (9) | NAFLD defined by the US-Fatty Liver Index (US-FLI) Score, using GGT | • Population: NHANES (1988-2016), participants aged >20.<br>• Sample size: 58,731 participants, (38.9% with Hypertension)<br>• Findings:<br>○ Higher risk of NAFLD in Hypertensive individuals (OR of 1.83 [95% CI 1.63-2.06]).<br>**Conclusion: There is a higher risk of NAFLD in Hypertensive individuals compared to normotensive individuals.** | No |
| Porepa 2010 (10) | No specific tests mentioned for NAFLD | • Population: Canadian general population (Ontario Health Insurance Plan database)<br>• Sample size: 2,497,777 participants, hypertensive participants = 352,732.<br>• Findings:<br>○ A positive association was found between diagnosed hypertension and a risk of serious liver disease (NAFLD or NASH, HR: 1.23 [95% CI 1.14–1.31]). | No |

| Source | Blood Tests | Evidence | Quality concerns |
|---|---|---|---|
| | | **Conclusion: Hypertensive individuals have a higher risk of NAFLD or NASH compared to normotensive individuals.** | |

#### 4.3 Is there anything the GP can do to manage or treat NAFLD?

No. Other than lifestyle advice (which is same recommends for patients with hypertension) there is not much the GP can do. This suggests there is no benefit to monitor HTN patients for NAFLD. Answered in the type 2 diabetes mellitus evidence report and presented in the Feb 2022 consensus meeting.

| Source | Evidence |
|---|---|
| NICE | Non-alcoholic fatty liver disease (NAFLD): assessment and management<br>NICE guideline [NG49]<br>Published: 06 July 2016<br>https://www.nice.org.uk/guidance/ng49/evidence/full-guideline-pdf-2548213310<br><br>• Weight reduction interventions: no relevant studies identified. Nevertheless, NICE recommends offering advice on physical activity and diet to people with NAFLD.<br>• Dietary modification and supplements: Do not offer omega-3 fatty acids to adults with NAFLD because there is not enough evidence to recommend their use.<br>• Exercise interventions: There is some evidence that exercise reduces liver fat content.<br>• Lifestyle modification: there is some evidence that lifestyle interventions (diet, behavioural modifications, and exercise) are beneficial (although the effect may be small). NICE recommends considering lifestyle interventions regardless of the patients’ BMI.<br>• Alcohol advice: Recommendation to keep alcohol consumption within national limits. More research is needed to determine whether reducing alcohol intake below national limits is beneficial for NAFLD patients.<br>• No evidence to make any recommendations on caffeine and fructose intake.<br>Pharmacological interventions: there is currently no licensed treatment for NAFLD. |

#### 5. **Lipid profile** (Serum cholesterol, High-density lipoprotein (HDL), Low-density lipoprotein (LDL), Triglycerides, HDL:LDL ratio)

##### 5.1 Which acute or chronic complications do these tests pick up?

Dyslipidaemia, which is a risk factor for CVD (NICE), including stroke and myocardial infarction, cerebrovascular disease, coronary heart disease, and ischemic heart disease (ICHOM). Lipid tests also used to calculate risk of CVD (QRISK 3 (11)) which uses total cholesterol and HDL cholesterol.

##### 5.2 Are dyslipidaemia and cardiovascular risk more common in HTN patients compared to the general population?

Hypertensive patients have higher mean values of Triglycerides and cholesterol compared to normotensive patients.
Hypertensive patients have a higher prevalence of CVD events.
Total cholesterol and HDL levels are used to calculate CVD risk, therefore there is good evidence to include them. However, unsure about LDL and HDL:LDL ratio as no evidence was found for those tests, but they can be calculated alongside total cholesterol and HDL levels.

| **Source** | **Blood Tests** | **Evidence** | **Quality concerns** |
|---|---|---|---|
| NICE Guidelines | total cholesterol and HDL cholesterol | For all people with hypertension offer to:<br>• take a blood sample to measure: total cholesterol and HDL cholesterol<br>https://www.nice.org.uk/guidance/ng136/resources/hypertension-in-adults-diagnosis-and-management-pdf-66141722710213 | |
| *Systematic reviews* | | | |
| Huang 2021 (12) | No specific tests mentioned for CVD | • Population: Patients with 2018 ESC guideline defined isolated diastolic hypertension and normotensive patients.<br>• Sample size: 15 studies (n = 489,814)<br>• Findings:<br> ○ Isolated diastolic hypertension was significantly associated with an increased risk of composite cardiovascular events (HR 1.28 [95% CI 1.07–1.52; p = 0.006]), cardiovascular mortality (HR 1.45 [95% CI: 1.07–1.95; p = 0.015]), all strokes (HR 1.44 [95% CI 1.04–2.01; p = 0.03]), and hemorrhagic stroke (HR 1.64 [95% CI 1.18–2.29; p = 0.164]).<br>**Conclusion: Hypertension increases the risk of cardiovascular events and strokes.** | No |
| Palla 2018 (13) | No specific tests mentioned for CVD | • Population: Patients treated for masked hypertension, white-coat hypertension, normotension, or sustained hypertension (SH).<br>• Sample size: 9 studies included, 14,729 participants (11,245 normotensives, 3,484 participants with masked hypertension, 1,984 participants with white-coat hypertension, and 5,143 participants with SH<br>• Findings:<br> ○ Cardiovascular events occurred in 12.3% of patients with masked hypertension and 5.1% of patients with normotension. The frequency of cardiovascular events was significantly higher in patients with masked hypertension (OR 2.91 [95% CI 2.54–3.33]).<br>**Conclusion: The prevalence of CVD is over 2 fold in hypertensive patients compared to normotensive patients.** | No mention of results of publication bias assessments. |

| Source | Blood Tests | Evidence | Quality concerns |
|---|---|---|---|
| Han 2020 (14) | No specific tests mentioned for CVD | • Population: Patients with stage 1 hypertension or normotension.<br>• Sample size: 11 articles (reporting on 16 independent studies, n = 212,447) with 65,945 CVD events.<br>• Findings:<br>o Risk of CVD events was increased in patients with stage 1 hypertension compared to normotensive patients (HR 1.38[ 95% CI 1.28–1.49]).<br>o The subgroup analyses found that stage 1 hypertension was associated with coronary heart disease (CHD) (HR 1.30 [95% CI 1.20–1.41]), stroke (HR 1.39 [95% CI 1.27–1.52]), CVD morbidity (HR 1.42, [95% CI 1.32–1.53]), and CVD mortality (HR 1.34 [95% CI 1.05–1.71]).<br>**Conclusion: The prevalence of CVD is higher in hypertensive patients compared to normotensive patients.** | No |
| *Primary Studies* | | | |
| Ren 2021 (15) | No specific tests mentioned for CVD | • Population: NHANES (2013-2018) participants aged 60+.<br>• Sample size: 4,346 subjects, stroke group: 182, Non-stroke group: 4,164<br>• Findings:<br>o Patients with hypertension are more likely to experience a stroke, (OR 2.3 [95% CI 1.34-3.94].<br>**Conclusion: those with hypertension have a higher risk of stroke.** | CVD defined by medical questionnaire. |
| Hua 2021 (3) | triglycerides (mg/dl), total cholesterol (mg/dl), | • Population: NHANES (2007-2018) participants aged 18-80.<br>• Sample size: 26,778, 49.6 of patents had hypertension.<br>• Findings: Mean values:<br>o Triglycerides (mg/dL):<br>▪ Normotensive: 106.0 (n = 13,495)<br>▪ Hypertensive: 138.0 (n = 13,283)<br>o Cholesterol (mg/dL):<br>▪ Normotensive: 187.0 (n = 13,495)<br>▪ Hypertensive: 194.0 (n = 13,283)<br>**Conclusion: Mean triglycerides and cholesterol values are higher in hypertensive patients compared to normotensive patients.** | No |

#### 5.3 Is there anything the GP can do to manage or treat this?

Yes, the GP can advise on diet and lifestyle changes, tight blood pressure control through lifestyle and antihypertensive drugs, and offering lipid modification therapy (usually statins).Answered in the type 2 diabetes mellitus evidence report and presented in the Feb 2022 consensus meeting.

| Source | Evidence |
|---|---|
| NICE | **Lipid modification - CVD prevention: Scenario: Lipid therapy - primary prevention of CVD**<br>Last revised in May 2021<br>https://cks.nice.org.uk/topics/lipid-modification-cvd-prevention/management/lipid-therapy-primary-prevention-of-cvd/<br>Consider offering lipid modification therapy (without the need for a formal risk assessment) to people who aged 85 years or over (particularly people who smoke or have raised blood pressure)<br>**Hypertension: Scenario: Management**<br>Last revised in July 2022<br>https://cks.nice.org.uk/topics/hypertension/management/management/<br>Discuss starting antihypertensive drug treatment, in addition to lifestyle advice, with people aged under 80 years with persistent stage 1 hypertension who have one or more of the following: target organ damage, established cardiovascular disease (CVD), renal disease, diabetes, an estimated 10-year risk of CVD of 10% or more.<br>Consider antihypertensive drug treatment in addition to lifestyle advice for people aged under 60 years with stage 1 hypertension and an estimated 10-year CVD risk below 10%. Bear in mind that 10-year cardiovascular risk may underestimate the lifetime probability of developing CVD. |

#### 5.4 Are there clear benefits of earlier detection or treatment?

Yes. Untreated elevated lipid levels increase the risk of CVD. Answered in the type 2 diabetes mellitus evidence report and presented in the Feb 2022 consensus meeting.

| Source | Evidence |
|---|---|
| NICE | **Lipid modification - CVD prevention:**<br>**What is the relationship between blood lipids and cardiovascular health?**<br>Last revised in May 2021<br>https://cks.nice.org.uk/topics/lipid-modification-cvd-prevention/background-information/lipids-cardiovascular-health/<br>"Total cholesterol is an important predictor of CVD events. However, non-high density lipoprotein cholesterol (non-HDL-C), the difference between total and HDL cholesterol, which usually makes up 60–70% of total serum cholesterol is a powerful risk factor for CVD."<br>"Elevated triglyceride levels (greater than 2.3 mmol/L), especially when HDL-C levels are low, is a risk factor for CVD and is independent of total cholesterol." "Extreme levels of triglycerides (greater than 20 mmol/L) are associated with pancreatitis and a high risk of morbidity and mortality." |

#### 6. Full blood count (haemoglobin)

##### 6.1 Which acute or chronic complications do these tests pick up?

Anaemia (low red blood cell count).

##### 6.2 Is anaemia more common in patients with hypertension compared to the general population?

No evidence was identified on the risk of anaemia amongst patients with hypertension.

| Source | Blood Tests | Evidence | Quality concerns |
|---|---|---|---|
| NICE Guidelines | Not Mentioned | | |
| *Systematic Reviews* | | | |
| None identified | | | |
| *Primary Studies* | | | |
| None identified | | | |

##### 6.3 Is there anything the GP can do to manage or treat this?

Yes, give dietary advice and for iron deficiency anaemia and prescribe iron tablets, for vitamin B12 deficiency anaemia refer or administer hydroxocobalamin, and for folate deficiency anaemia prescribe folic acid. Answered in the type 2 diabetes mellitus evidence report and presented in the Feb 2022 consensus meeting.

| Source | Evidence |
|---|---|
| NICE | **Anaemia - iron deficiency: Management (CKS)**<br>Last revised in November 2021<br>https://cks.nice.org.uk/topics/anaemia-iron-deficiency/management/<br>Iron deficiency anaemia:<br>• Prescribe all people with iron deficiency anaemia one tablet once daily of oral ferrous sulfate, ferrous fumarate or ferrous gluconate — continue treatment for 3 months after iron deficiency is corrected to allow stores to be replenished.<br>• Monitor the person to ensure that there is an adequate response to iron treatment.<br><br>**Anaemia - B12 and folate deficiency (CKS)**<br>Last revised in July 2020<br>https://cks.nice.org.uk/topics/anaemia-b12-folate-deficiency/<br>Vit B12 deficiency anaemia:<br>• For people with neurological involvement: Seek urgent specialist advice from a haematologist.<br>• For people with no neurological involvement: Initially administer hydroxocobalamin 1 mg intramuscularly three times a week for 2 weeks. The maintenance dose depends on whether the deficiency is diet related or not. Give dietary advice about foods that are a good source of vitamin B12.<br>Folate deficiency anaemia: |

| Source | Evidence |
| --- | --- |
| | • Prescribe oral folic acid 5 mg daily — in most people, treatment will be required for 4 months.<br>• Check vitamin B12 levels in all people before starting folic acid.<br>• Give dietary advice about foods that are a good source of folic acid. |

#### 6.4 Are there clear benefits of earlier detection or treatment?

Yes, untreated anaemia can worsen long term patient outcomes (more evidence or expert input needed). For instance, iron deficiency in patients with low ejection fraction is associated with decreased long-term survival and lower quality of life. Answered in the type 2 diabetes mellitus evidence report and presented in the Feb 2022 consensus meeting.

| Source | Evidence | Quality Concerns |
| --- | --- | --- |
| Wienbergen 2018 (16) | • Population: Germany and Switzerland patients with heart failure with reduced ejection fraction<br>• Sample size: 949 patients (Iron deficiency (n = 505) without iron deficiency (n = 418)<br>• Findings:<br>○ Patients with iron deficiency had a higher long-term mortality compared to those without iron deficiency (19.5% vs. 13.7%, p = 0.02) and reported a lower quality of life. Only a minority of patients with ID (9.3%) received iron supplementation during long-term course, just 4.7% intravenously.<br>○ In the adjusted analysis a significant interaction remained, with iron deficiency being a significant predictor of 1-year mortality in patients without anaemia (HR 2.15, 95% CI 1.12–3.78), but not in anaemic patients (HR 0.99, 95% CI 0.65–1.49).<br>**Conclusion: Anaemia was associated with an elevated mortality.** | No |

#### 7. **Haematinics** (Vit B12, Serum ferritin, Folate tests)

##### 7.1 Which acute or chronic complications do these tests pick up?

Vitamin B12, ferritin, and folate deficiencies, also called iron deficiency anaemia, vitamin B12 and folate anaemia.

##### 7.2 Is anaemia, or vitamin B12, ferritin, and folate deficiencies more common in patients with hypertension compared to the general population?

No evidence was identified on the risk of vitamin B12, ferritin, and folate deficiencies amongst patients with hypertension.

| **Source** | **Blood Tests** | **Evidence** | **Quality concerns** |
|---|---|---|---|
| NICE Guidelines | Not mentioned | | |
| *Systematic Reviews* | | | |
| None identified | | | |
| *Primary Studies* | | | |
| None identified | | | |

##### 7.3 Is there anything the GP can do to manage or treat this?

See answer to 6.3

##### 7.4 Are there clear benefits of earlier detection or treatment?

See answer to 6.4

#### 8. Thyroid function tests (Thyroxine, thyroid stimulating hormone, Free triiodothyronine (T3), free thyroxine, serum T3 level)

##### 8.1 Which acute or chronic complications do these tests pick up?

Hyper- or hypothyroidism.

##### 8.2 Are hyper- or hypothyroidism more common in patients with hypertension compared to the general population?

No evidence was identified on the risk of hyperthyroidism or hypothyroidism amongst patients with hypertension.

| Source | Blood Tests | Evidence | Quality concerns |
|---|---|---|---|
| NICE Guidelines | Not mentioned | | |
| *Systematic Reviews* | | | |
| None identified | | | |
| *Primary Studies* | | | |
| None identified | | | |

##### 8.3 Is there anything the GP can do to manage or treat hyper- or hypothyroidism?

Yes, the GP can prescribe pharmacological treatments, such as levothyroxine for primary or subclinical hypothyroidism, or radioactive iodine for adults with Graves' disease or hyperthyroidism secondary to multiple nodules. Answered in the type 2 diabetes mellitus evidence report and presented in the Feb 2022 consensus meeting.

| Source | Evidence |
|---|---|
| NICE | Thyroid disease: assessment and management<br>NICE guideline [NG145]; Published: 20 November 2019<br>https://www.nice.org.uk/guidance/ng145<br>- Offer levothyroxine as first-line treatment for adults, children and young people with primary hypothyroidism.<br>- Consider levothyroxine for adults with subclinical hypothyroidism who have a TSH of 10 mlU/litre or higher on 2 separate occasions 3 months apart. Follow the recommendations in section 1.4 on follow-up and monitoring of hypothyroidism.<br>- Offer radioactive iodine as first-line definitive treatment for adults with Graves' disease, unless antithyroid drugs are likely to achieve remission (see recommendation 1.6.11), or it is unsuitable (for example, there are concerns about compression, malignancy is suspected, they are pregnant or trying to become pregnant or father a child within the next 4 to 6 months, or they have active thyroid eye disease).<br>- Offer radioactive iodine as first-line definitive treatment for adults with hyperthyroidism secondary to multiple nodules unless it is unsuitable (for example, there are concerns about compression, thyroid malignancy is suspected, they are pregnant or trying to become pregnant or father a child within the next 4 to 6 months, or they have active thyroid eye disease). |

##### 8.4 Are there clear benefits of earlier detection or treatment?

More evidence or expert input needed.

#### 9. **Clotting tests** (Prothrombin time, Partial thromboplastin time, Thrombin time)

##### 9.1 Which acute or chronic complications do these tests pick up?

Bleeding disorders.

##### 9.2 Are bleeding disorders or abnormal clotting test results more common in HTN patients compared to the general population?

No evidence was identified on the risk of bleeding disorders amongst HTN patients.

| **Source** | **Blood Tests** | **Evidence** | **Quality concerns** |
|---|---|---|---|
| NICE Guidelines | Not mentioned | | |
| *Systematic Reviews* | | | |
| None identified | | | |
| *Primary Studies* | | | |
| None identified | | | |

##### 9.3 Is there anything the GP can do to manage or treat this?

(more evidence or expert input needed).

| **Source** | **Evidence** |
|---|---|
| NICE | Platelets - abnormal counts and cancer:<br>Scenario: Management of platelet count outside the normal range (CKS)<br>Last revised in June 2021<br>https://cks.nice.org.uk/topics/platelets-abnormal-counts-cancer/management/management/<br>- The GP needs to establish the cause underlying the bleeding problems in order to treat them. |

##### 9.4 Are there clear benefits of earlier detection or treatment?

(more evidence or expert input needed)

#### 10. Bone profile (Serum inorganic phosphate, calcium, calcium adjusted, vitamin D, alkaline phosphatase, parathyroid hormone)

##### 10.1 Which acute or chronic complications do these tests pick up?

Serum calcium and phosphorus levels, vitamin D deficiency, parathyroid problems

##### 10.2 Are bone cancer, vitamin D deficiency, or parathyroid problems (or abnormal bone profile results) more common in patients with hypertension compared to the general population?

One study suggests that vitamin D levels are lower in hypertensive patients compared to normotensive patients.
No evidence was identified on the risk of parathyroid problems or serum calcium and phosphorus levels in patients with hypertension.

| Source | Blood Tests | Evidence | Quality concerns |
|---|---|---|---|
| NICE Guidelines | Not mentioned | | |
| *Systematic Reviews* | | | |
| None Identified | | | |
| *Primary Studies* | | | |
| Forrest 2011 (17) | Serum Vitamin D | • Population: NHANES, participants aged >20.<br>• Sample size: 4,495 subjects, vitamin D deficiency n = 2,257, no vitamin D deficiency n = 2,238<br>• Findings:<br>○ 1,482 subjects (30%) had hypertension. Those with vitamin D deficiency were more likely to have hypertension (33.5% [95% CI 30.6-36.5]) compared to patients without vitamin D deficiency (27.6 [95% CI 25.0-30.3]).<br>**Conclusion: There is an association between hypertension and vitamin D deficiency.** | Results in reverse, study does not take into account which condition is diagnosed first, so data is compatible for our report. |

#### 10.3 Is there anything the GP can do to manage or treat this

Yes, treatments are available for vitamin D deficiency, phosphate levels, and hyperparathyroidism (and calcium levels through hyperparathyroidism management).

Answered in the type 2 diabetes mellitus evidence report and presented in the Feb 2022 consensus meeting.

| Source | Evidence |
|---|---|
| NICE | **Vitamin D deficiency in adults:**<br>**Scenario: Management of vitamin D deficiency or insufficiency (CKS)**<br>Last revised in September 2021<br>https://cks.nice.org.uk/topics/vitamin-d-deficiency-in-adults/management/management/<br>• Advise that oral vitamin D3 is the vitamin D preparation of choice for the treatment of vitamin D deficiency, and vitamin D2 is an alternative option in some clinical situations.<br><br>**Hyperparathyroidism (primary): diagnosis, assessment and initial management**<br>NICE guideline [NG132]<br>Published: 23 May 2019<br>https://www.nice.org.uk/guidance/ng132<br>• Surgical management<br>• Non-surgical management if surgery is unsuccessful.<br><br>**Hyperphosphataemia in chronic kidney disease**<br>**NICE clinical guideline 157, part of managing chronic kidney disease (CG182) and managing anaemia in CKD (NG8)**<br>Published: March 2013<br>**https://www.nice.org.uk/guidance/ng203/evidence/full-guideline-2013-hyperphosphataemia-in-ckd-pdf-9206080240**<br>• Adults with hyperphosphataemia in chronic kidney disease should be offered a calcium-based phosphate binder as a first-line treatment in addition to dietary management |

#### 10.4 Are there clear benefits of earlier detection or treatment?

(more evidence or expert input needed)

#### 11. B-type natriuretic peptide (BNP)

##### 11.1 Which acute or chronic complications do these tests pick up?

Heart failure (NICE, CKS).

##### 11.2 Is heart failure more common in T2DM patients compared to the general population?

One study found that hypertension was associated with heart failure.

| Source | Blood Tests | Evidence | Quality concerns |
|---|---|---|---|
| NICE Guidelines | High blood pressure (hypertension) is one of the most important, treatable causes of premature morbidity and mortality in the world. It is a major risk factor for heart failure. https://www.nice.org.uk/guidance/ng136/resources/hypertension-in-adults-diagnosis-and-management-pdf-66141722710213 | | |
| *Systematic Reviews* | | | |
| None Identified | | | |
| *Primary Studies* | | | |
| Johansson 2001 (18) | | • Population: UK population, General Practice Research Database (GPRD)<br>• Sample size: 938 cases of heart failure, 5000 controls (matched on age and sex).<br>• Findings:<br>○ 1449/5000 (29.0%) of the controls without incident heart failure had hypertension, whereas 398/938 (42.4%) of the cases had hypertension also (RR 1.7 [95% CI 1.4 - 2.0]).<br>**Conclusion: there is an association between incident heart failure and hypertension.** | Results reversed. Study does not take into account which condition is diagnosed first, so data is compatible for our report |

#### 11.3 Is there anything the GP can do to manage or treat this?

Yes, there are treatments available for heart failure. Answered in the type 2 diabetes mellitus evidence report and presented in the Feb 2022 consensus meeting.

| Source | Evidence |
|---|---|
| NICE | **Heart failure - chronic:**<br>**Scenario: Confirmed heart failure with reduced ejection fraction (CKS)**<br>Last revised in August 2021<br>https://cks.nice.org.uk/topics/heart-failure-chronic/management/confirmed-heart-failure-with-reduced-ejection-fraction/<br>For a person with confirmed heart failure and reduced ejection fraction:<br>- Ensure drugs which may cause or worsen heart failure are reviewed and stopped if appropriate.<br>- To relieve symptoms of fluid overload, ensure the person has been prescribed a loop diuretic.<br>- Prescribe both an angiotensin-converting enzyme (ACE) inhibitor and a beta-blocker licensed to treat heart failure but only start one drug at a time.<br>For a person with confirmed heart failure with preserved ejection fraction:<br>- Ensure drugs which may cause or worsen heart failure are reviewed and stopped if appropriate.<br>- Prescribe a loop diuretic — up to 80 mg furosemide (or equivalent), if necessary, to relieve symptoms of fluid overload.<br>- Refer the person to a specialist for further advice on management.<br>- Consider if an antiplatelet drug is indicated.<br>- Consider prescribing an antiplatelet to people with atherosclerotic arterial disease (including coronary heart disease).<br>- Consider if statin therapy is indicated.<br>- Ensure that any causes, comorbidities, and precipitating factors are optimally managed.<br>- People with heart failure due to valve disease should be referred for specialist assessment and advice regarding follow-up. |

#### 11.4 Are there clear benefits of earlier detection or treatment?

(more evidence or expert input needed)

#### Tests to screen for adverse treatment effects

- Treatments considered:
  - ACE-inhibitors (ACEs), Angiotensin II receptor blockers (ARB), calcium channel blockers (CCB), and Thiazide type diuretic (TTDs).
- Adverse treatment effects considered:
  - Those mentioned in BNF:
    - Electrolyte imbalance, renal impairment, hyperkalemia – common or uncommon (most drugs listed this side effect)
    - Hepatic disorders, hepatitis and abnormal hepatic function – rare or very rare (most drugs listed this side effect)
    - Anaemia – rare or very rare (some drugs listed this side effect)
  - Blood glucose and new onset diabetes – Not listed in BNF but evidence was found during the rapid reviews
- Corresponding lab tests considered:
  - Blood glucose (any blood glucose monitoring test as HbA1c may not be used in a trial setting)
  - Renal function tests (eGFR, blood urea, blood electrolytes)
  - Liver function tests
  - Anaemia tests (Full blood count, iron, vitamin B12 and foliate blood tests)

### 1. Blood Glucose

#### 1.1 Is increased blood glucose or new onset diabetes more common in people who take antihypertensive drugs?

There is a risk for both increased blood glucose and new onset diabetes in patients that take TTDs, no risk was found for those on CCBs and a decrease in risk for both increased blood glucose and new onset diabetes in patients on ACEs and ARBs.

| Source | Blood Tests | Evidence | Quality concerns |
|---|---|---|---|
| BNF | Not mentioned | | |
| *Systematic Reviews* | | | |
| Nazarzadeh 2021 (19) | No specific blood test mentioned | Drug Groups: **ACEIs, ARBs, CCBs and TTDs**<br>• Population: individual participant data from Blood Pressure Lowering Treatment Trialists' Collaboration (BPLTTC).<br>• Sample size: 22 included trials (n = 145,939), consisting of 19 randomised controlled trials, 8 placebo-controlled, and 14 drug class comparison trials.<br>• Findings:<br> ○ ACEIs (RR 0.84 [95% CI 0.76–0.93]) and ARBs (RR 0.84 [95% CI 0.76–0.92]) both reduced the risk of type 2 diabetes when compared with placebo.<br> ○ CCBs did not affect the risk of type 2 diabetes when compared with placebo (RR 1.02 [95% CI 0.92–1.13]).<br> ○ Thiazide diuretics were found to increase the risk of type 2 diabetes compared with placebo (RR 1.20 [95% CI 1.07–1.35]).<br>**Conclusion: Thiazide diuretics increase the risk of new onset diabetes, CCBs have no effect on the risk of type 2 diabetes, and ACEIs and ARBs reduced the risk of new onset diabetes.** | Most of the trials included were drug class comparison trials, which reduced the drug vs placebo sample size. |
| Li 2017 (20) | no specific blood test mentioned | Drug groups: **ACEs, ARBs, CCBs and TTDs**<br>• Population: Hypertensive patients without diabetes<br>• Sample size: 38 studies (patients on antihypertensives (n = 224,140), vs on placebo (n = 58,174))<br>• Findings:<br> ○ ACEIs (OR 0.82 [95% CI 0.66–1.00]) and ARBs (OR 0.77 [95% CI 0.62–0.93]) had no effect on the risk of developing new onset diabetes compared to placebo.<br> ○ CCBs had no effect on risk of developing new onset diabetes compared to placebo (OR: 1.00 [95% CI 0.82-1.30]).<br> ○ TTDs increased the risk of developing new onset diabetes compared to placebo (OR: 1.40 [95% CI 1.10-1.80]).<br>**Conclusion: Diuretics increase the risk of new onset diabetes, CCBs have no effect on the risk of type 2 diabetes, and ACEIs and ARBs reduced the risk of new onset diabetes.** | Type of diuretic not specified in review, most primary studies included use of chlorthalidone, which is a thiazide type diuretic, so this review is reported with the assumption that the diuretic used is chlorthalidone. Heterogeneity not addressed, and no mention of sensitivity analyses. |

| Source | Blood Tests | Evidence | Quality concerns |
|---|---|---|---|
| Zhang 2016 (21) | Fasting plasma glucose (FPG) | Drug groups: **TTDs**<br>• Population: Hypertensive patients without diabetes<br>• Sample size: 26 studies, (n = 16,162)<br>• Findings:<br>○ Thiazide or thiazide-type diuretics were found to increase Fasting Plasma Glucose level compared with non-thiazide agents or placebo or nontreatment (Mean difference (MD): 0.27 mmol/L (4.86 mg/dL) [95% CI 0.15–0.39]).<br>○ In a further analysis, where non-thiazide agents were removed, thiazide-type diuretics did not significantly increase FPG level when compared with placebo or nontreatment (thiazide vs placebo or nontreatment (MD: 0.21 mmol/L (3.78 mg/dL) [95% CI 0.00–0.41]).<br>**Conclusion: Thiazide-type diuretics do not significantly increase FPG levels in hypertensive patients, but when compared to non-thiazide drugs (ACEs, ARBs and CCBs), Thiazide type diuretics may seem to increase FPG due to non- thiazide drugs potential glucose level lowering effects.** | Potential insufficient search strategy, and unclear in methods if studies were reviewed by 2 independent reviewers. |
| *Primary Studies* | | | |
| None Identified | | | |

#### 1.2 Is there anything the GP can do to manage or treat increased blood glucose or new onset diabetes?

Yes, other than stopping or changing medication and dosage, there are several options for medications which can be changed in dose or combination to control blood glucose levels. Answered in the type 2 diabetes mellitus evidence report and presented in the Feb 2022 consensus meeting.

#### 1.3 Are there clear benefits of earlier detection or treatment?

Yes, early intervention with drugs has long term benefits through lowering blood glucose and reducing the complications of Diabetes. Answered in the type 2 diabetes mellitus evidence report and presented in the Feb 2022 consensus meeting.

### 2. Renal function tests

#### 2.1 Is abnormal renal function more common in those who take hypertensive drugs?

ACE inhibitors increase the risk of hyperkalaemia, however there is conflicting evidence on how ACEs affect renal function.
ARBs increase the risk of acute renal injury and hyperkalaemia.
No evidence was found on the effects of CCBs and TTDs on renal function.

| Source | Blood Tests | Evidence | Quality concerns |
|---|---|---|---|
| BNF | Electrolyte imbalance, renal impairment and hyperkalaemia.<br>Drug groups ACEs, ARBs and TTDs listed this as common (1 in 100 to 1 in 10) or uncommon (1 in 1000 to 1 in 100) side effects. | | |
| *Systematic reviews* | | | |
| Takuathung 2022 (22) | No specific blood test mentioned | Drug groups: **ACEs**<br>• Population: General population<br>• Sample size: 257 studies included (18 studies reporting on hyperkalemia (n = 12,471), 19 studies reporting on renal impairment (n = 85,864)).<br>• Findings:<br> ○ Compared with a placebo, ACE inhibitors were significantly associated with an increased risk hyperkalaemia (RR: 1.24 [95% CI 1.01-1.52]).<br> ○ Compared with a placebo, ACE inhibitors were not associated with an increased risk in renal impairment (RR: 1.13 [95% CI 0.72-1.75]).<br>**Conclusion: ACEs increase the risk of hyperkalaemia and does not increase risk of renal impairment.** | No |
| Bavishi 2016 (23) | No specific blood test mentioned | Drug groups: **ACEs**<br>• Population: Patients on ACEIs, placebo or active controls, aged >65 either with or at high risk for CVD.<br>• Sample size: 16 studies, (n = 104,321 (4 studies assessing ACEs vs placebo (n = 4,838), 5 studies assessing ACEs vs active controls (n = 62,665).<br>• Findings:<br> ○ ACEIs were associated with an increased risk of renal insufficiency (RR 1.29 [95% CI 1.02-1.63]) compared with placebo.<br> ○ ACEIs were associated with a decreased risk of renal insufficiency (RR 0.87 [95% CI 0.70 -1.07]) compared with active control.<br>**Conclusion: Conflicting evidence for risk of renal insufficiency for ACEs.** | No information of risk of bias being addressed for synthesis, and only older patients included. |
| Elgendy 2015 (24) | serum creatinine<br>serum potassium levels | Drug groups: **ARBs**<br>• Population: Patients aged >65.<br>• Sample size: 16 studies in total (n = 113,386), 8 trials with placebos, (n= 50,522, 25,101 patients in the ARB group and 25,421 in the placebo group.)<br>• Findings:<br> ○ ARBs increase the risk of acute kidney injury (RR 1.61 [95% CI 1.27–2.04]) compared with placebo.<br> ○ ARBs increase the risk of hyperkalaemia (RR 2.12 [95% CI 1.61–2.80]) compared with placebo. | Search conducted on Medline only, and only older patients included. 8 of the 16 trials used other drugs as controls, reducing the sample size. |

| Source | Blood Tests | Evidence | Quality concerns |
| --- | --- | --- | --- |
| | | **Conclusion: ARBs increase the risk of acute renal injury and hyperkalaemia.** | |
| *Primary studies* | | | |
| None identified | | | |

#### 2.2 Is there anything the GP can do to manage or treat increased abnormal renal function or electrolyte imbalance?

Yes, hyperkalaemia can be treated by changing medication and dosage and lifestyle changes.

#### 2.3 Are there clear benefits of earlier detection or treatment?

Yes, early intervention can halt further kidney damage and rising potassium levels by antihypertensive drugs.

### 3. Liver function tests

#### 3.1 Is hepatitis or abnormal liver function more common in people who take antihypertensive drugs?

No evidence was found on Hepatic disorders, hepatitis and abnormal hepatic function as a side effect of hypertensive drugs.

| Source | Blood Tests | Evidence | Quality concerns |
|---|---|---|---|
| BNF | Hepatic disorders, hepatitis and abnormal hepatic function.<br>- Drug groups ACEs and ARBs listed these as rare (1 in 10 000 to 1 in 1000) or very rare (<1 in 10 000) side effects.<br>Some specific drugs within the TTD and CCBS drug groups listed these as rare (1 in 10 000 to 1 in 1000), very rare (<1 in 10 000) or frequency unknown side effects. | | |
| *Systematic Reviews* | | | |
| None identified | | | |
| *Primary Studies* | | | |
| None identified | | | |

#### 3.2 Is there anything the GP can do to manage or treat increased abnormal liver function?

Other than stopping or changing medication and dosage and lifestyle changes there is not much that can be done.

#### 3.3 Are there clear benefits of earlier detection or treatment?

Yes, early intervention can halt further liver damage by antihypertensive drugs .

### 4. Anaemia

#### 4.1 Is Anaemia more common in people who take antihypertensive drugs?

No evidence was found on Anaemia as a side effect of hypertensive drugs.

| Source | Blood Tests | Evidence | Quality concerns |
|---|---|---|---|
| BNF | Anaemia<br>- Drug groups ACEs and TTDs listed this as a rare (1 in 10 000 to 1 in 1000) or very rare (<1 in 10 000) side effect.<br>Some specific drugs within the ARBs drug group listed this as a rare (1 in 10 000 to 1 in 1000), very rare (<1 in 10 000) side effect. | | |
| *Systematic Reviews* | | | |
| None identified | | | |
| *Primary Studies* | | | |
| None identified | | | |

#### 4.2 Is there anything the GP can do to manage or treat Anaemia?

Yes, see answer to 6.3

#### 4.3 Are there clear benefits of earlier detection or treatment?

Yes, early intervention can halt further lowering of iron, folate, vitamin B12, and haemoglobin levels.

#### Abbreviations

| | |
|---|---|
| ACE | Angiotensin-converting enzyme |
| ALP | Alkaline phosphatase |
| ALT | Alanine aminotransferase |
| AST | Aspartate transaminase |
| BNF | British National Formulary |
| BNP | Brain Natriuretic Peptide |
| CGM | Continuous Glucose Monitoring |
| CKD | Chronic Kidney Disease |
| CRP | C-reactive protein |
| CVD | Cardiovascular disease |
| eGFR | Estimated Glomerular Filtration Rate |
| ESR | Erythrocyte Sedimentation Rate |
| ESRD | End-stage renal disease |
| FPG | Fasting plasma glucose |
| GA | Glycated albumin |
| HbA1c | Glycated haemoglobin |
| HDL | High-density lipoprotein |
| HF | Heart failure |
| HTN | Hypertension |
| ICHOM | International Consortium for Health Outcomes Measurement |
| KNHANES | Korean National Health and Nutrition Examination Survey |
| LDL | Low-density lipoprotein |
| MI | Myocardial infarction |
| NAFLD | Non-alcoholic fatty liver disease |
| NHANES | National Health and Nutrition Examination Survey |
| NICE | National Institute for Health and Care Excellence |
| PVD | Peripheral Vascular Disease |
| RRT | Renal Replacement Therapy |
| T1DM | Type 1 diabetes mellitus |
| T2DM | Type 2 diabetes mellitus |
| UKPDS | UK Prospective Diabetes Study |
