## Supplementary Tables and Figures for "Evidence-based blood tests for monitoring adults with hypertension in primary care: rapid review, routine data analyses, and consensus study"

Supplementary materials


#### Authors

Martha MC Elwenspoek,[1,2] Rachel O'Donnell,[2] Catalina Lopez Manzano,[2] Sarah Dawson,[1] Lewis Buss,[2] Katie Charlwood,[2] Christina Stokes,[3] Francesco Palma,[3] Alastair D Hay,[2] Penny Whiting,[2] Jessica Watson.[2]

#### Affiliations

1. The National Institute for Health Research Applied Research Collaboration West (NIHR ARC West), University Hospitals Bristol NHS Foundation Trust, BS1 2NT Bristol, UK
2. Population Health Sciences, Bristol Medical School, University of Bristol, BS8 2PS Bristol, UK
3. Patient representative

#### Corresponding author

Martha M C Elwenspoek
9th Floor, Whitefriars, Lewins Mead,
Bristol, BS1 2NT

Tel: +44/0 117 3427689

### Search strategies systematic reviews

#### Table S1. Rapid review 1: How prevalent are the following acute or chronic complications in hypertensive patients compared to the general population?

Including: CKD, Heart failure, Type 2 diabetes, CVD, liver disease, thyroid, bleeding disorders, and bone profile related conditions.

**Database: KSR Evidence**

| | Terms | Hits |
|---|---|---|
| #1 Hypertension | hypertension or hyper-tension or ("blood pressure*" AND (primary or essential or high* or elevated or abnormal* or monitor*)) or "HTN" or hyperpiesi* or hypertensive* or hyper-tensive* or (*hypertensi** OR *hyper-tensi** OR ("*blood pressure*"* NEAR (*primary* OR *essential* OR *high** OR *elevated* OR *abnormal** OR monitor*))) | 7705 |
| #2 Prevalence | prevalence or incidence or predict* or risk* | 114453 |
| #3 Chronic kidney disease | CKD or “kidney function*” or “kidney failure*” or “renal function*” or “renal failure*” or nephropathy or nephropathies or nephrologic* or glomerular filtration rate*” or (kidney AND (disorder* or disease* or dysfunction* or chronic* or insufficienc* or impair*)) or (renal AND (disorder* or disease* or dysfunction* chronic* or insufficienc* or impair*)) or (“kidney failure*” NOT (graft* or allograft* or transplant*)) not (renal NEAR (dialysis or surgery or haemodialysis or hemodialysis or transplant)) not (kidney NEAR (dialysis or surgery or haemodialysis or hemodialysis or transplant)) | 6116 |
| #4 | #1 AND #3 | 938 |
| #5 | #2 AND #4 | 758 |
| #6 Heart failure | “Heart failure” or “cardiac failure” | 3264 |
| #7 | #1 AND #6 | 597 |
| #8 | #2 AND #7 | 488 |
| #9 Type 2 diabetes | ("diabetes type 2" or "diabetes type II" or "type 2 diabet*" or "type II diabet*" or (diabet* near ("type 2" or "type II")) or “diabetes mellitus type 2” or “diabetes mellitus type II” or T2DM or "T2 DM" or DMT2 or "DM T2" or "adult onset diabet*" or "late onset diabet*" or "matur* onset diabet*" or "NIDDM" or "non insulin* diabet*" or "noninsulin* diabet*" or "insulin independent diabet*" or "ketosis resistant diabet*") | 5114 |
| #10 | #1 AND #9 | 729 |
| #11 | #2 AND #10 | 585 |
| #12 Cardiovascular disease | “myocardial infarction” or stroke* or “brain ischemia” or “cerebrovascular accident” or (cardiac NEAR (death or sudden or mortality)) or apoplex* or “Abnormal heart rhythm*” or arrhythmia* or “marfan syndrome” or “ Deep vein thrombosis” or “pulmonary embolism” or “heart attack” or cardiomyopathy or “intracranial h?emorrhage*” or “coronary syndrome” or (cardiovascular NEAR (event* or mortality or death*)) or (disease* or disorder* NEAR (cardiovascular or coronary or heart or artery or cardiac or aorta or “Congenital heart” or “Coronary artery” or “Heart muscle” or “Heart valve” or pericardial or “Peripheral vascular” or “Rheumatic heart” or Vascular or “blood vessel” or “isch?emic heart” or cerebrovascular)) | 67317 |

| | | |
|---|---|---|
| #13 | #1 AND #12 | 4389 |
| #14 | #2 AND #13 | 3583 |
| #15 Liver disease | “NAFLD” or “liver function” or “fatty liver” or “cirrhosis” or “cirrho*” or “steatohepatitis” or “fibrosis” or “fibro*” or “non-alcoholic steatohepatitis” or “NASH” or “hepatic*” or “hepato*” or (“hepatic” AND (“complications” or “co-morbid*”)) | 8104 |
| #16 | #1 AND #15 | 435 |
| #17 | #2 AND #16 | 322 |
| #18 Anaemia | “anaem*” or “anem*” or “iron-poor blood” or “Low blood” or “Tired blood” or “iron” or “ferritin” or “folate” or “folic acid” or “vitamin B9” or “B12” or “cobalamin” or “deficien*” or “total iron” or “iron level*” or “levels of iron” or “iron store*” or “stored iron” or “heme iron” or “haem iron” or ((“vitamin” or “iron” or “ferritin” or “folate” or “folic acid” or “vitamin B9” or “B12” or “cobalamin”) AND (“deficien*” or “low” or “insufficien*” or “lack” or “levels”)) | 5968 |
| #19 | #1 AND #18 | 379 |
| #20 | #2 AND #19 | 301 |
| #21 bone profile | “parathyroid*” or “hypoparathyroidism” or “hyperparathyroidism” or “PTH” or “parathormone” or “parathyrin” or “Pseudohypoparathyroidism” or ((bone or bones or 3artilage* or chondro* or osteo* or spine or spinal or skeletal or extraskeletal or Ewing) AND (cancer* or carcinoma* or neoplas* or metast* or sarcoma* or tumo?r*)) or chondrosarcoma* or “vitamin D” or “vitamin D deficien*” or “insufficien* vitamin D” or “Cholecalciferol” or “ergocalciferol” or ((“vitamin D” ) AND (“deficien*” or “low” or “insufficien*” or “lack” or “levels”)) | 4047 |
| #22 | #1 AND #21 | 152 |
| #23 | #2 AND #22 | 118 |
| #24 thyroid disorders | “Thyroid disease” or “thyroid disorders” or “thyroid dysfunction” or “dysfunctional thyroid” or “hypothyroidism” or “underactive thyroid” or “hyperthyroidism” or “hyper-thyroid*” or “overactive thyroid” or “Hashimoto’s” or “Hashimotos” or “thyroiditis” or “Graves’” or “Graves” or “thyroxin*” or “thyroid stimulating hormone” | 861 |
| #25 | #1 AND #24 | 53 |
| #26 | #2 AND #25 | 44 |
| #27 | (bleeding or platelet AND (disorder* or problem*)) or "platelet function defects" or "Disseminated intravascular coagulation" or DIC or "Prothrombin deficiency" or h?emophilia or "Glanzmann disease" or "Idiopathic thrombocytopenic purpura" or ITP or "Von Willebrand disease" or (factor AND deficiency) | 6422 |
| #28 | #1 AND #24 | 318 |
| #29 | #2 AND #25 | 267 |

#### Table S2. Rapid review 2: How prevalent are abnormal test results in hypertensive patients compared to the general population?

Including: Renal function tests, Lipid profile tests, Natriuretic Peptide Tests, HbA1c

**Database: KSR Evidence**

| Search component | Terms | Hits |
|---|---|---|
| #1 Hypertension | hypertension or hyper-tension or ("blood pressure*" AND (primary or essential or high* or elevated or abnormal* or monitor*)) or "HTN" or hyperpiesi* or hypertensive* or hyper-tensive* or (*hypertensi** OR *hyper-* | 7708 |

| | | |
|---|---|---|
| | *tensi** OR ("*blood pressure**" NEAR (*primary* OR *essential* OR *high** OR *elevated* OR *abnormal** OR monitor*))) | |
| #2 Prevalence | Prevalence or prevalence studies or incidence or prediction or predict* or risk* | 114561 |
| #3 Renal function tests | "Serum creatinine" or potassium or sodium or electrolyte* or "Urea blood" or "nitrate blood" or "Blood urea nitrogen" or "urine protein" or BUN or GFR or eGFR or "Glomerular filtration rate" or "albumin: creatinine ratio" or "albumin creatinine" ratio or ACR or Urinalysis | 4320 |
| #4 | #1 AND #3 | 545 |
| #5 | #2 AND #4 | 404 |
| #6 Lipid profile tests | "Serum cholesterol" or "total cholesterol" or" High density lipoprotein" or HDL or "Low density lipoprotein" or LDL or Triglyceride* or "HDL:LDL ratio" or "HDL LDL ratio" or "Lipid Profile" or "Fasting Lipid*" or "Non fasting Lipid*" or "Cholesterol Test*" | 2955 |
| #7 | #1 AND #6 | 875 |
| #8 | #2 AND #7 | 715 |
| #9 Natriuretic Peptide Tests | "Natriuretic Peptide Tests" or "BNP" or "NT-proBNP" or "brain natriuretic peptide" or "N-terminal pro b-type natriuretic peptide" or "B-type natriuretic peptide" or "brain natriuretic factor" or "BNF" | 390 |
| #10 | #1 AND #9 | 58 |
| #11 | #2 AND #10 | 41 |
| #12 Hba1c | HbA1c or "h?emoglobin A1*" or "glycated h?emoglobin" or "glycated ha?emoglobin" or "glycosylated Hb" or GHb or "plasma glucose" or "total A1*" or glycoh?emoglobin or HgbA1c or Hb1c | 2317 |
| #13 | #1 AND #12 | 411 |
| #14 | #2 AND #13 | 312 |
| #15 Clotting tests | "Clotting test*" or "Coagulation Test*" or "Prothrombin time" or "Partial thromboplastin time" or "Thrombin time" or INR or "Factor V assay" or "Fibrinogen level" or "PT" or "PT-INR" or "Platelet count" | 853 |
| #16 | #1 AND #15 | 32 |
| #17 | #2 AND #16 | 27 |

#### Table S3. Rapid review 3: Hypertensive Drugs and the side effects

Side effects for hypertensive drugs that can monitored by blood tests including liver disease and liver function tests, kidney disease and kidney function tests, anaemia, lipid tests, and heart failure tests.

**Database: KSR Evidence**

| | Terms | |
|---|---|---|
| #1 | "hypertensive drug*" or "hypertension drug*" or "Angiotensin converting enzyme inhibitor*" or ACE* or "Angiotensin II receptor blocker*" or ARB* or AIIRA* or "Thiazide type diuretic*" or TTD* or "Calcium channel blocker*" or CCB* or Captopril or Enalapril or Fosinopril or Imidapril or Lisinopril or "Perindopril arginine" or "Perindopril erbumine" or Quinapril or Ramipril or Trandolapril or Azilsartan or candesartan or Eprosartan or Irbesartan or Losartan or Olmesartan or Telmisartan or Valsartan or Chlortalidone or Indapamide or metolazone or xipamide or hydrochlorothiazide or Bendroflumethiazide or Amlodipine or Felodipine or Nifedipine or Diltiazem or Lacidipine or Lercanidipine or Nicardipine or Nimodipine or Verapamil | 4978 |
| #2 | hypertension or hyper-tension or ("blood pressure*" AND (primary or essential or high* or elevated or abnormal* or monitor*)) or "HTN" or hyperpiesi* or hypertensive* or hyper-tensive* or (hypertensi* OR hyper-tensi* OR ("blood | 8242 |

| | | |
|---|---|---|
| | pressure*" NEAR (primary OR essential OR high* OR elevated OR abnormal* OR monitor*))) NOT hypotension | |
| #3 | #1 AND #2 | 643 |
| #4 | “Serum creatinine” or potassium or sodium or electrolyte* or “Urea blood” or “nitrate blood” or “Blood urea nitrogen” or “urine protein” or BUN or GFR or eGFR or “Glomerular filtration rate” or “albumin: creatinine ratio” or “albumin creatinine” ratio or ACR or Urinalysis or CKD or “Electrolyte imbalance” or hyperkalaemia or “kidney function*” or “kidney failure*” or “renal function*” or “renal failure*” or nephropathy or nephropathies or nephrologic* or glomerular filtration rate*” or (kidney AND (disorder* or disease* or dysfunction* or chronic* or insufficienc* or impair*)) or (renal AND (disorder* or disease* or dysfunction* chronic* or insufficienc* or impair*)) or (“kidney failure*” NOT (graft* or allograft* or transplant*)) not (renal NEAR (dialysis or surgery or haemodialysis or hemodialysis or transplant)) not (kidney NEAR (dialysis or surgery or haemodialysis or hemodialysis or transplant)) | 9628 |
| #5 | #3 AND #4 | 185 |
| #6 | “liver function test*” or “Albumin” or “Alkaline phosphatase” or “ALP” or “Alanine aminotransferase” or “Alanine transaminase” or “ALT” or “Aspartate aminotransferase” or “Aspartate transaminase” or “AST” or “Bilirubin” or “Gamma glutamyl transpeptidase” or “Gamma-glutamyltransferase” or “GGT” or “Total protein” or “Serum globulin” or “Liver enzymes” or “Albumin and total protein” or “L-lactate dehydrogenase” or “LD” or “Prothrombin time” or “PT” or NAFLD or “liver function” or “fatty liver” or cirrhosis or cirrho* or steatohepatitis or fibrosis or fibro* or “non-alcoholic steatohepatitis” or NASH or hepatic* or hepato* or (hepatic AND (disorders or complications or co-morbid*)) | 10848 |
| #7 | #3 AND #6 | 55 |
| #8 | “anaem*” or “anem*” or “Aplastic an?emia” or “Haemolytic an?emia” or “hypoplastic an?emia” or “iron-poor blood” or “Low blood” or “Tired blood” or “iron” or “ferritin” or “folate” or “folic acid” or “cobalamin” or “deficien*” or “total iron” or “iron level*” or “levels of iron” or “iron store*” or “stored iron” or “heme iron” or “haem iron” or ((“vitamin” or “iron” or “ferritin” or “folate” or “folic acid” or “cobalamin”) AND (“deficien*” or “low” or “insufficien*” or “lack” or “levels”)) | 6332 |
| #9 | #3 AND #8 | 26 |
| #10 | “Serum cholesterol” or “total cholesterol” or” High density lipoprotein” or HDL or “Low density lipoprotein” or LDL or Triglyceride* or “HDL:LDL ratio” or “HDL LDL ratio” or “Lipid Profile” or “Fasting Lipid*” or “Non fasting Lipid*” or “Cholesterol Test*” | 3200 |
| #11 | #3 AND #10 | 35 |
| #12 | “Natriuretic Peptide Tests” or “BNP” or “NT-proBNP” or “brain natriuretic peptide” or “N-terminal pro b-type natriuretic peptide” or “B-type natriuretic peptide” or “brain natriuretic factor” or “BNF” | 419 |
| #13 | #3 AND #12 | 2 |

### Search strategies primary research

#### Table S4. Rapid review 4: Combined search

- Rapid Review 1a: How prevalent are these acute or chronic complications in hypertensive patients compared to the general population?
- Rapid review 1b: How prevalent are abnormal test results in hypertensive patients compared to the general population?

**Database: EMBASE (1974 to 2022 June 14) and MEDLINE (1946 to June 14, 2022)**

| Nr | Search component | Search terms | Hits |
|---|---|---|---|
| 1 | Hypertension | hypertension or hyper-tension or ("blood pressure*" AND (high* or elevated or abnormal*)) or HTN or hyperpiesi* or hypertensive* or hyper-tensive* or (hypertensi* OR hyper-tensi* OR ("blood pressure*" NEAR (high* OR elevated OR abnormal*))) | 1670603 |
| 2 | Primary care/health care databases; UK/European registries; cohorts | (CRPD or Clinical Practice Research Data* or (GOLD adj3 (data* or regist*)) or AURUM or General Practice Research Data* or GPRD or Health Improvement Network* or THIN data* or QResearch or Q-Research or NHANES or (National Health adj2 Nutrition Examination Survey) or (Information System? adj3 (Development and Research and Primary Care)) or SIDIAP or Caserta or BIFAP or Base de Datos para la Investigacion Farmacoepidemiologica en Atencion Primaria or ((Finnish or Danish or Swedish) adj3 (national or regional or health*) adj3 (registr* or register)) or DNPR or ((electronic Data Research adj2 Innovation Service) or eDRIS) or (Integrated Primary Care Information Data* or IPCI) or (((Quintiles or IMS) adj Disease Analys*) or mediplus) or ((Quintiles* or IMS or LPD) adj3 health adj3 data*) or ((Quintiles* or IMS) adj3 (longitudinal or patient) adj3 data*) or ((Norwegian or Polish or Czech or MODY) adj3 (Registr* or register)) or Pedianet Data* or (Securite Sociale adj2 Maladie) or (German adj2 (Pharmacoepidemiolog* or Pharmaco-epidemiolog*) adj3 Data*) or Secure Anonymi#ed Information Link* or (ALSPAC or (Avon longitudinal adj3 (cohort or study))) or National Child Development Study or British Cohort Study or (Next Steps adj3 data*)).mp. | 78451 |
| 3 | | 1 and 2 | 8875 |
| 4 | Animal studies | ((animal model* or mouse or mice or murine* or rat or rats or rodent* or muridae or murids or rabbit* or leporine* or leporidae or guineapig* or cavies or caviidae or hamster* or cricetidae or gerbil* or gerbillinae or cat or cats or feline* or felidae or dog or dogs or canine* or canidae or pig or pigs or piglet* or minipig* or swine* or porcine* or suidae or horse or horses or donkey or donkies or burros or equine* or equidae or sheep or lamb or lambs or ovine or ovidae or goat or goats or cow or cows or cattle or bovine* or bovidae or primate* or monkey or monkeys or macaque or macaques or marmoset or marmosets) not human*).ti. | 4840028 |
| 5 | Pregnancy | (((gestational or maternal* or pregnan*) adj5 hypertensi*) or eclamp* or pre-eclamp* or preeclamp*).ti,kf,hw. | 131709 |
| 6 | Pregnancy | (((gestational or maternal* or pregnan*) adj3 complication?) and (cardio* or hypertensi*)).ti,kf,hw. | 30553 |

| | | | |
|---|---|---|---|
| 7 | Pregnancy | ((gestational or maternal* or pregnan*) and (HELLP syndrom* or h?emolysis elevated liver enzymes low platelet count*)).ti,kf,hw. | 7311 |
| 8 | Covid | (covid or covid19 or covid-19 or covid2019 or covid-2019 or ncov* or novel coronavirus or novel betacoronavirus or sars-ncov-2 or sars-cov-2 or postcovid* or longcovid*).ti,kf,hw. | 499583 |
| 9 | Children | ((infant* or child* or schoolchild*) not adult*).ti. | 2098060 |
| 10 | | 4 or 5 or 6 or 7 or 8 or 9 | 7548518 |
| 11 | | 3 not 10 | 8303 |
| 12 | | (serum creatinin? or (blood? adj1 (urea or nitrate? or nitrite?)) or ((urine or urea) adj3 protein?) or proteinuria? or GFR or eGFR or glomerular filtration rate? or albumin?creatinin? ratio or (albumin? adj3 creatinin? adj3 ratio?) or urinalysis or urinalyses or albuminuria?).mp. | 761788 |
| 13 | | ((potassium or sodium or electrolyte?) and (renal* or kidney* or function test*)).mp. | 251641 |
| 14 | | ((potassium or sodium or electrolyte?) adj2 (test or tests or testing)).mp. | 3213 |
| 15 | Renal function tests | 12 or 13 or 14 | 962608 |
| 16 | | (CKD or CKF or nephropath* or nephritis or nephrologic* or ((renal* or kidney*) adj3 (disorder* or disease* or dysfunction* or chronic* or failure* or insufficienc* or impair*))).mp. | 1281475 |
| 17 | | (graft* or allograft* or transplant*).ti,kf,hw. | 1927933 |
| 18 | | (ESRD or ((endstage? or end-stage?) adj3 (kidney* or renal*) adj3 (disease* or dysfunction* or failure* or impairment* or insufficienc*))).ti,kf,hw. | 76650 |
| 19 | | (h?emodialys* or ((renal* or kidney* or extracorporeal or extra-corporeal or blood?) adj (dialysis or dialyses))).ti,kf,hw. | 268496 |
| 20 | Chronic kidney disease | 16 not (17 or 18 or 19) | 946954 |
| 21 | | 15 or 20 | 1636676 |
| 22 | | 11 and 21 | 1698 |
| 23 | | remove duplicates from 22 | 1205 |
| 24 | | (HbA1c or Hb-A1c or h?emoglobin A1* or glyc* h?emoglobin or glyc* Hb or GHb or total A1* or glycoh?emoglobin or HgbA1c or Hgb-A1c or Hb1c or Hb-1c).mp. | 248791 |
| 25 | | plasma glucose.mp. | 89074 |
| 26 | | ((diabet* adj3 (type-2 or type II)) or T2DM or T2 DM or DMT2 or DM T2 or adult onset diabet* or late onset diabet* or matur* onset diabet* or NIDDM or non insulin* diabet* or noninsulin* diabet* or insulin independent diabet* or ketosis resistant diabet*).mp. | 510016 |
| 27 | | diabet*.ti. | 901472 |
| 28 | Diabetes | 24 or 25 or 26 or 27 | 1196662 |
| 29 | | 11 and 28 | 1598 |
| 30 | | remove duplicates from 29 | 1105 |
| 31 | | (clot* test* or coagulat* test* or co-agulat* test* or prothrombin time? or pro-thrombin time? or partial thromboplastin time? or partial thrombo-plastin time? or thrombin time? or INR or PT-INR or factor V assay? or fibrinogen level? or platelet? count*).mp. | 285818 |
| 32 | | (((bleeding or platelet?) and (disorder* or problem*)) or platelet? function defects or disseminated intravascular coagulation or prothrombin deficien* or pro-thrombin deficien* or h?emophilia* or Glanzmann disease or idiopathic thrombocytopenic purpura or idiopathic thrombo-cytopenic purpura or ITP or Von Willebrand disease).mp. | 319841 |
| 33 | | (factor? and deficienc*).mp. | 356175 |

| | | | |
|---|---|---|---|
| 34 | Clotting tests | 31 or 32 or 33 | 881989 |
| 35 | | 11 and 34 | 176 |
| 36 | | remove duplicates from 35 | 139 |
| 37 | BNF | (natriuretic peptide? or BNP or NT-proBNP or NT-pro-BNP or brain natriuretic factor? or BNF).mp. | 129782 |
| 38 | | 11 and 37 | 17 |
| 39 | | ((cardiac or heart or myocardial) adj failure?).mp. | 708425 |
| 40 | | 11 and 39 | 786 |
| 41 | | 38 or 40 | 793 |
| 42 | | remove duplicates from 41 | 609 |
| 43 | | (NAFLD or liver function or fatty liver or cirrho* or steatohepatitis or steato-hepatitis or NASH or ((liver or hepatic or hepato*) and (fibro* or complication? or comorbid* or co-morbid*))).mp. | 1078187 |
| 44 | | liver disease?.ti,kf,hw. | 312324 |
| 45 | NAFLD | 43 or 44 | 1190874 |
| 46 | | 11 and 45 | 351 |
| 47 | | remove duplicates from 46 | 282 |
| 48 | | (an?em* or low blood? or tired blood? or ((iron or ferritin or folate or folic acid or B9 or vitaminB9 or B12 or vitaminB12 or cobalamin*) and (deficien* or low* or insufficien* or lack* or level?))).mp. | 972051 |
| 49 | | (iron or ferritin or folate or folic acid or B9 or vitaminB9 or B12 or vitaminB12 or cobalamin*).ti,kf. | 302885 |
| 50 | | 48 or 49 | 1100861 |
| 51 | | 11 and 50 | 342 |
| 52 | | remove duplicates from 51 | 240 |
| 53 | | (parathyroid* or para-thyroid* or hypoparathyroid* or hypo-parathyroidi* or hypo-para-thyroid* or hyperparathyroid* or hyper-parathyroid* or hyper-para-thyroid*or PTH or parathormon* or para-thormon* or parathyrin* or para-thyrin* or pseudohypoparathyroidism or ((bone or bones or cartilage* or chondro* or osteo* or spine or spinal or skeletal or extraskeletal or Ewing) and (cancer* or carcinoma* or neoplas* or metast* or sarcoma* or tumo?r*)) or chondrosarcoma*).mp. | 1164005 |
| 54 | | ((vitamin D or vitaminD or cholecalciferol or ergocalciferol) and (deficien* or low* or insufficien* or lack* or level?)).mp. | 166072 |
| 55 | | (vitamin D or vitaminD or cholecalciferol or ergocalciferol).ti,kf. | 108111 |
| 56 | Bone profile | 53 or 54 or 55 | 1308356 |
| 57 | | 11 and 56 | 321 |
| 58 | | remove duplicates from 57 | 228 |
| 59 | Thyroid function | (thyroid* or hypothyroid* or hypo-thyroid* or hyperthyroidism or hyper-thyroid* or Hashimoto* or Graves* or thyroxin*).mp. | 687565 |
| 60 | | 11 and 59 | 160 |
| 61 | | remove duplicates from 60 | 121 |
| 62 | | (23 or 30 or 36 or 42 or 47 or 52 or 58 or 61) | 2766 |
| 63 | | remove duplicates from 62 | 2741 |

#### Table S5. Rapid review 5: Hypertension drug side effects

Side effects for hypertensive drugs reported in known randomised controlled trials.

**Database: EMBASE (1974 to 2022 September 22) and MEDLINE (1946 to September 22, 2022)**

| Nr | Search terms | Hits |
|---|---|---|
| 1 | (adverse or complication* or drug induced or harm or harms or health risks or potential risks or negative effects or noxious or safe* or side effect* or side reaction* or tolerance or tolerated or tolerabilit* or toxicity or toxicities).tw,kf. | 8246413 |
| 2 | ((undesirable or harm* or serious or toxic) adj3 (effect? or reaction? or event? or outcome?)).tw,kf. | 358194 |
| 3 | Special Situation for Pharmacovigilance.fs. | 191399 |
| 4 | Unexpected Outcome of Drug Treatment.fs. | 34163 |
| 5 | (ae or si or to or co).fs. | 7753068 |
| 6 | adverse.ox. | 433903 |
| 7 | exp adverse drug reaction/ | 723562 |
| 8 | exp drug toxicity/ | 276011 |
| 9 | exp intoxication/ | 398641 |
| 10 | exp drug safety/ | 513919 |
| 11 | exp drug monitoring/ | 81665 |
| 12 | exp drug hypersensitivity/ | 110534 |
| 13 | exp postmarketing surveillance/ | 38195 |
| 14 | exp drug surveillance program/ | 26631 |
| 15 | exp phase iv clinical trial/ | 7310 |
| 16 | or/3-15 | 8985867 |
| 17 | 16 use oemezd | 4631729 |
| 18 | exp product surveillance, postmarketing/ | 55915 |
| 19 | exp adverse drug reaction reporting systems/ | 12480 |
| 20 | Drug Hypersensitivity/ | 77796 |
| 21 | exp drug monitoring/ | 81665 |
| 22 | exp Drug Toxicity/ | 276011 |
| 23 | exp poisoning/ | 565495 |
| 24 | (ae or co or ci or de or po or to).fs. | 10783730 |
| 25 | Clinical trial phase IV.pt. | 2362 |
| 26 | or/18-25 | 11347042 |
| 27 | 26 use medall | 7069428 |
| 28 | 1 or 2 or 17 or 27 | 16797484 |
| 29 | risk.ti. | 1308284 |
| 30 | 28 or 29 | 17648591 |
| [BPLTTC Trials] | | |
| 31 | ALLHAT*.af. | 971 |
| 32 | NCT00000542.af. | 75 |
| 33 | "antihypertensive and lipid lowering treatment to prevent heart attack*".af. | 489 |
| 34 | or/31-33 | 1038 |
| 35 | 28 and 34 | 527 |
| 36 | remove duplicates from 35 | 381 |
| 37 | (ANBP? adj2 (cohort or study or trial or RCT or randomi* controlled trial)).af. | 70 |
| 38 | "Australian National Blood Pressure Study".af. | 138 |

| | | |
|---|---|---|
| 39 | 37 or 38 | 155 |
| 40 | 30 and 39 | 67 |
| 41 | remove duplicates from 40 | 48 |
| 42 | ASCOT-BPLA.af. | 116 |
| 43 | (Anglo-Scandinavian Cardiac Outcomes Trial and (BPLA or ((BP or Blood Pressure) adj Lowering Arm))).af. | 72 |
| 44 | 42 or 43 | 140 |
| 45 | 30 and 44 | 85 |
| 46 | remove duplicates from 45 | 60 |
| 47 | (CAMELOT and Amlodipine and Enalapril).af. | 21 |
| 48 | (Amlodipine and Enalapril and Limit and Occurrence? and Thrombo*).af. | 14 |
| 49 | 47 or 48 | 26 |
| 50 | remove duplicates from 49 | 16 |
| 51 | Captopril Prevention Project.af. | 75 |
| 52 | (CAPPP adj2 (cohort or study or trial or RCT or randomi* controlled trial)).af. | 39 |
| 53 | 51 or 52 | 85 |
| 54 | 30 and 53 | 48 |
| 55 | remove duplicates from 54 | 32 |
| 56 | CARDIO-SIS.af. | 14 |
| 57 | CARDIOvascolari del Controllo della Pressione Arteriosa SIStolica.af. | 7 |
| 58 | 56 or 57 | 14 |
| 59 | remove duplicates from 58 | 11 |
| 60 | (CASE-J adj2 (cohort or study or trial or RCT or randomi* controlled trial)).af. | 64 |
| 61 | (Case-J and candesartan).af. | 64 |
| 62 | (Candesartan Antihypertensive Survival Evaluation adj2 Japan).af. | 45 |
| 63 | or/60-62 | 80 |
| 64 | 30 and 63 | 54 |
| 65 | remove duplicates from 64 | 35 |
| 66 | (COLM adj2 (cohort or study or trial or RCT or randomi* controlled trial)).af. | 14 |
| 67 | NCT00454662.af. | 2 |
| 68 | (Combin* adj2 OLMesartan adj2 (CCB or Calcium Channel Blocker?) adj5 Diuretic?).af. | 5 |
| 69 | (OLMesartan and (CCB or Calcium Channel Blocker?) and diuretic? and elderly and hypertens*).af. | 35 |
| 70 | or/66-69 | 41 |
| 71 | remove duplicates from 70 | 30 |
| 72 | (Controlled Onset Verapamil Investigation adj2 Cardiovascular End Points).af. | 8 |
| 73 | ((CONVINCE adj2 (cohort or study or trial or RCT or randomi* controlled trial)) and verapamil).af. | 25 |
| 74 | 72 or 73 | 30 |
| 75 | remove duplicates from 74 | 16 |
| 76 | "Combination Therapy of Hypertension to Prevent Cardiovascular Events".af. | 20 |
| 77 | (NCT00135551 or UMIN000001152).af. | 8 |
| 78 | ((COPE adj2 (cohort or study or trial or RCT or randomi* controlled trial)) and benidipine).af. | 22 |
| 79 | or/76-78 | 24 |

| | | |
|---|---|---|
| 80 | remove duplicates from 79 | 14 |
| 81 | "Dutch Transient Isch?emic Attack Trial".af. | 2 |
| 82 | "Dutch TIA Trial".af. | 54 |
| 83 | ((Dutch TIA adj2 (cohort or study or trial or RCT or randomi* controlled trial)) and aspirin).af. | 37 |
| 84 | or/81-83 | 61 |
| 85 | remove duplicates from 84 | 40 |
| 86 | "Efficacy of Candesartan on Outcome in Saitama Trial".af. | 2 |
| 87 | (ECOST and (Candesartan or hypertens*)).af. | 1 |
| 88 | (Candesartan and (Saitama adj2 (cohort or study or trial or RCT or randomi* controlled))).af. | 3 |
| 89 | (hypertens* and (Saitama adj2 (cohort or study or trial or RCT or randomi* controlled))).af. | 6 |
| 90 | (Candesartan and Saitama and hypertens*).af. | 55 |
| 91 | 30 and 90 | 36 |
| 92 | or/86-89,91 | 41 |
| 93 | remove duplicates from 92 | 34 |
| 94 | ((Lacidipine adj2 (cohort or study or trial or RCT or randomi* controlled)) and Atherosclerosis).af. | 83 |
| 95 | ((ELSA adj2 (cohort or study or trial or RCT or randomi* controlled)) and Lacidipine and (Atherosclerosis or hypertens*)).af. | 34 |
| 96 | 94 or 95 | 93 |
| 97 | 30 and 96 | 36 |
| 98 | remove duplicates from 97 | 27 |
| 99 | "European trial on reduction of cardiac events with perindopril in stable coronary artery".af. | 54 |
| 100 | ((EUROPA adj2 (cohort or study or trial or RCT or randomi* controlled)) and perindopril).af. | 126 |
| 101 | ISRCTN37166280.af. | 4 |
| 102 | or/99-101 | 161 |
| 103 | 30 and 102 | 90 |
| 104 | remove duplicates from 103 | 63 |
| 105 | EWPHE.af. | 85 |
| 106 | "European Working Party on High Blood Pressure in the Elderly".af. | 101 |
| 107 | 105 or 106 | 133 |
| 108 | 30 and 107 | 102 |
| 109 | remove duplicates from 108 | 69 |
| 110 | HDFP.af. | 190 |
| 111 | "Hypertension Detection and Follow-up Program".af. | 315 |
| 112 | 110 or 111 | 380 |
| 113 | 30 and 112 | 160 |
| 114 | remove duplicates from 113 | 106 |
| 115 | HOMED-BP.af. | 55 |
| 116 | (Hypertens* and (Objective Treatment adj3 Measur* adj3 Electrical Device? adj3 Blood Pressure)).af. | 39 |
| 117 | 115 or 116 | 55 |
| 118 | remove duplicates from 117 | 34 |
| 119 | "Heart Outcomes Prevention Evaluation Study".af. | 77 |

| | | |
|---|---|---|
| 120 | ((HOPE adj2 (cohort or study or trial or RCT or randomi* controlled)) and ramipril and (cardiovascular adj (risk? or event?))).af. | 215 |
| 121 | 119 or 120 | 263 |
| 122 | 30 and 121 | 195 |
| 123 | remove duplicates from 122 | 138 |
| 124 | "Hypertension in the Very Elderly Trial".af. | 157 |
| 125 | (HYVET adj2 (cohort or study or trial or RCT or randomi* controlled)).af. | 156 |
| 126 | NCT00122811.af. | 24 |
| 127 | 124 or 125 or 126 | 214 |
| 128 | 127 and 30 | 132 |
| 129 | remove duplicates from 128 | 93 |
| 130 | "International Nifedipine GITS Study".af. | 53 |
| 131 | "Intervention as a Goal in Hypertension Treatment".af. | 68 |
| 132 | ((INSIGHT adj2 (cohort or study or trial or RCT or randomi* controlled)) and Nifedipine).af. | 60 |
| 133 | or/130-132 | 108 |
| 134 | 30 and 133 | 58 |
| 135 | remove duplicates from 134 | 39 |
| 136 | (International Verapamil adj2 Trandolapril Study).af. | 182 |
| 137 | ((INVEST adj2 (cohort or study or trial or RCT or randomi* controlled)) and Verapamil and Trandolapril).af. | 150 |
| 138 | NCT00133692.af. | 39 |
| 139 | or/136-138 | 209 |
| 140 | 30 and 139 | 146 |
| 141 | remove duplicates from 140 | 97 |
| 142 | JMIC-B.af. | 18 |
| 143 | "Japan Multicenter Investigation for Cardiovascular Diseases-B".af. | 13 |
| 144 | 142 or 143 | 20 |
| 145 | remove duplicates from 144 | 13 |
| 146 | "Losartan Intervention for Endpoint Reduction in Hypertension".af. | 303 |
| 147 | 146 and 30 | 181 |
| 148 | remove duplicates from 147 | 132 |
| 149 | (National Intervention Cooperative Study adj2 Elderly Hypertens*).af. | 7 |
| 150 | (NICS-EH and hypertens*).af. | 12 |
| 151 | 149 or 150 | 17 |
| 152 | remove duplicates from 151 | 11 |
| 153 | (NORDIL and Diltiazem).af. | 64 |
| 154 | "Nordic Diltiazem Study".af. | 33 |
| 155 | 153 or 154 | 73 |
| 156 | 155 and 30 | 42 |
| 157 | remove duplicates from 156 | 32 |
| 158 | "Ongoing Telmisartan Alone and in Combination with Ramipril Global Endpoint Trial".af. | 247 |
| 159 | ((ONTARGET adj2 (cohort or study or trial or RCT or randomi* controlled)) and Telmisartan and Ramipril).af. | 333 |
| 160 | 158 or 159 | 381 |

| | | |
|---|---|---|
| 161 | 160 and 30 | 317 |
| 162 | remove duplicates from 161 | 206 |
| 163 | (prevent* and Atherosclerosis and Ramipril and Auckland).af. | 30 |
| 164 | (Prevention adj3 Atherosclerosis adj3 Ramipril).af. | 5 |
| 165 | 163 or 164 | 35 |
| 166 | remove duplicates from 165 | 35 |
| 167 | "Prevention of Events with Angiotensin Converting Enzyme Inhibition".af. | 53 |
| 168 | ((PEACE adj2 (cohort or study or trial or RCT or randomi* controlled)) and angiotensin converting enzyme inhibit*).af. | 57 |
| 169 | NCT00000558.af. | 12 |
| 170 | or/167-169 | 82 |
| 171 | 30 and 170 | 45 |
| 172 | remove duplicates from 171 | 33 |
| 173 | (Prospective Randomized Evaluation adj3 Vascular Effects adj3 (Norvasc or amlodipine)).af. | 31 |
| 174 | ((PREVENT adj2 (cohort or study or trial or RCT or randomi* controlled)) and (Norvase or amlodipine)).af. | 51 |
| 175 | 173 or 174 | 55 |
| 176 | remove duplicates from 175 | 35 |
| 177 | "Perindopril Protection Against Recurrent Stroke".af. | 175 |
| 178 | ((PROGRESS adj (cohort or study or trial or RCT or randomi* controlled)) and Perindopril).af. | 101 |
| 179 | 177 or 178 | 244 |
| 180 | 30 and 179 | 163 |
| 181 | remove duplicates from 180 | 115 |
| 182 | "Systolic Hypertension in the Elderly Program".af. | 343 |
| 183 | ((SHEP adj2 (cohort or study or trial or RCT or randomi* controlled)) and hypertens*).af. | 123 |
| 184 | 182 or 183 | 391 |
| 185 | 30 and 184 | 221 |
| 186 | remove duplicates from 185 | 144 |
| 187 | Systolic Blood Pressure Intervention Trial.af. | 890 |
| 188 | (SPRINT Trial and (Systolic Blood Pressure or SBP)).af. | 272 |
| 189 | NCT01206062.af. | 202 |
| 190 | or/187-189 | 1077 |
| 191 | 30 and 190 | 651 |
| 192 | remove duplicates from 191 | 457 |
| 193 | "STOP HYPERTENSION-2".af. | 48 |
| 194 | "Swedish Trial in Old Patients with Hypertension-2".af. | 27 |
| 195 | 193 or 194 | 62 |
| 196 | remove duplicates from 195 | 33 |
| 197 | (SYST-EUR adj2 (cohort or study or trial or RCT or randomi* controlled)).af. | 256 |
| 198 | (Systolic Hypertension adj2 Europe adj2 (cohort or study or trial or RCT or randomi* controlled)).af. | 70 |
| 199 | 197 or 198 | 294 |
| 200 | 30 and 199 | 187 |
| 201 | remove duplicates from 200 | 128 |

| | | |
|---|---|---|
| 202 | "Telmisartan Randomised Assessment Study in ACE Intolerant Subjects with Cardiovascular Disease".af. | 19 |
| 203 | ((TRANSCEND adj2 (cohort or study or trial or RCT or randomi* controlled)) and Telmisartan).af. | 60 |
| 204 | 202 or 203 | 74 |
| 205 | 30 and 204 | 54 |
| 206 | remove duplicates from 205 | 35 |
| 207 | "Valsartan in Elderly Isolated Systolic Hypertension".af. | 7 |
| 208 | (Valsartan and Systolic Hypertens* and elderly).af. | 98 |
| 209 | 30 and 208 | 84 |
| 210 | VALISH.af. | 18 |
| 211 | NCT00151229.af. | 4 |
| 212 | 207 or 209 or 210 or 211 | 98 |
| 213 | remove duplicates from 212 | 73 |
| 214 | "Valsartan Antihypertensive Long-Term Use Evaluation".af. | 108 |
| 215 | ((VALUE adj (cohort or study or trial or RCT or randomi* controlled)) and (Valsartan and (follow* up or followup or long* term or longterm))).af. | 92 |
| 216 | 214 or 215 | 128 |
| 217 | 30 and 216 | 97 |
| 218 | remove duplicates from 217 | 65 |
| 219 | "Verapamil in Hypertension and Atherosclerosis Study".af. | 14 |
| 220 | (VHAS and Verapamil and Hypertens*).af. | 17 |
| 221 | 219 or 220 | 21 |
| 222 | remove duplicates from 221 | 15 |
| 223 | (Blood Pressure Lowering Treatment Trial* Collaboration or BPLTTC).af. | 91 |
| 224 | 30 and 223 | 53 |
| 225 | remove duplicates from 224 | 39 |
| 226 | 36 or 41 or 46 or 50 or 55 or 59 or 65 or 71 or 75 or 80 or 85 or 93 or 98 or 104 or 109 or 114 or 118 or 123 or 129 or 135 or 141 or 145 or 148 or 152 or 157 or 162 or 166 or 172 or 176 or 181 or 186 or 192 or 196 or 201 or 206 or 213 or 218 or 222 or 225 | 2730 |
| 227 | remove duplicates from 226 | 2720 |

### PRISMA flow diagrams

Figure S1. Rapid review 1: How prevalent are the following acute or chronic complications in hypertensive patients compared to the general population?

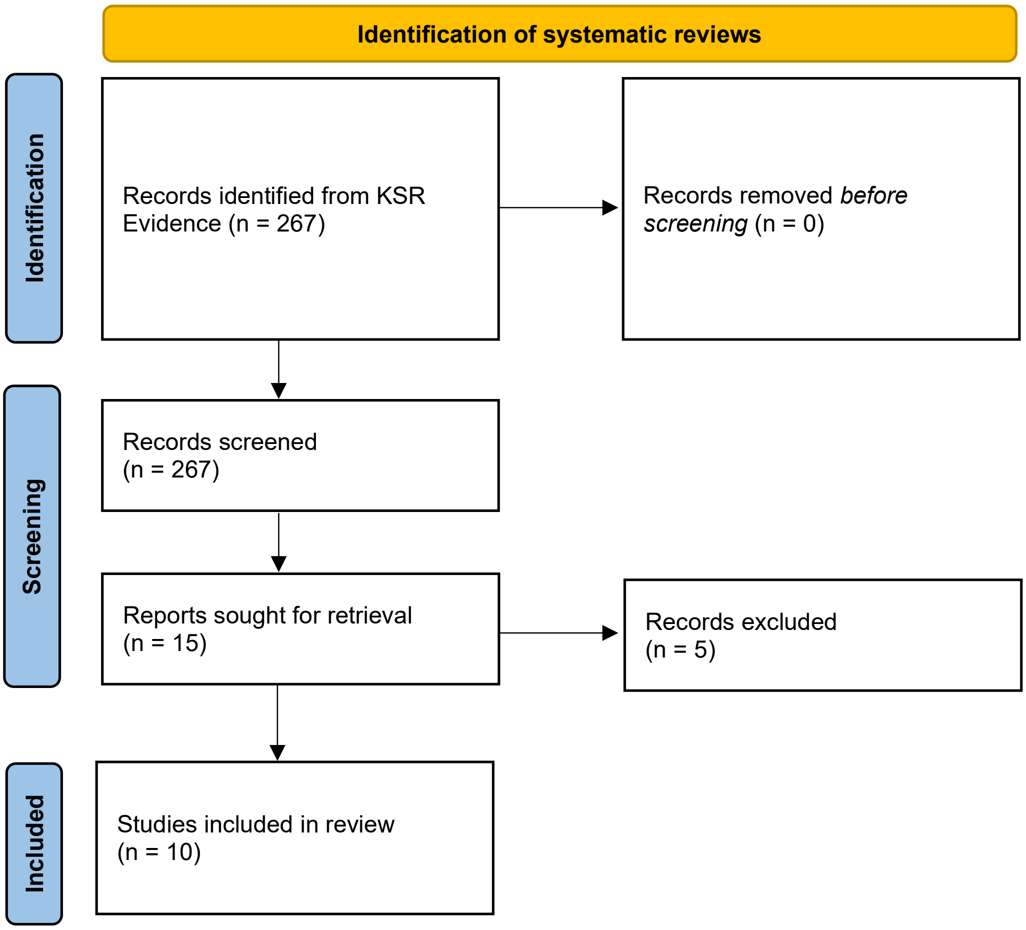

Figure S2. Rapid review 2: How prevalent are abnormal test results in hypertensive patients compared to the general population?

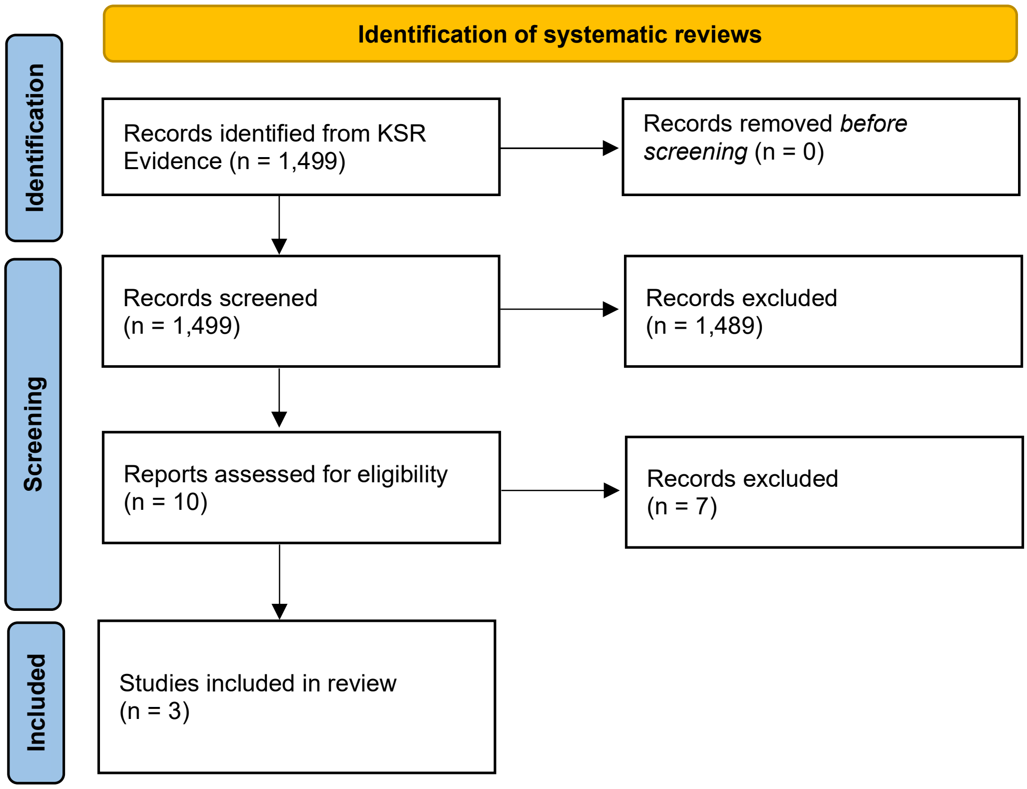


Figure S3. Rapid review 3: Hypertensive Drugs and the side effects

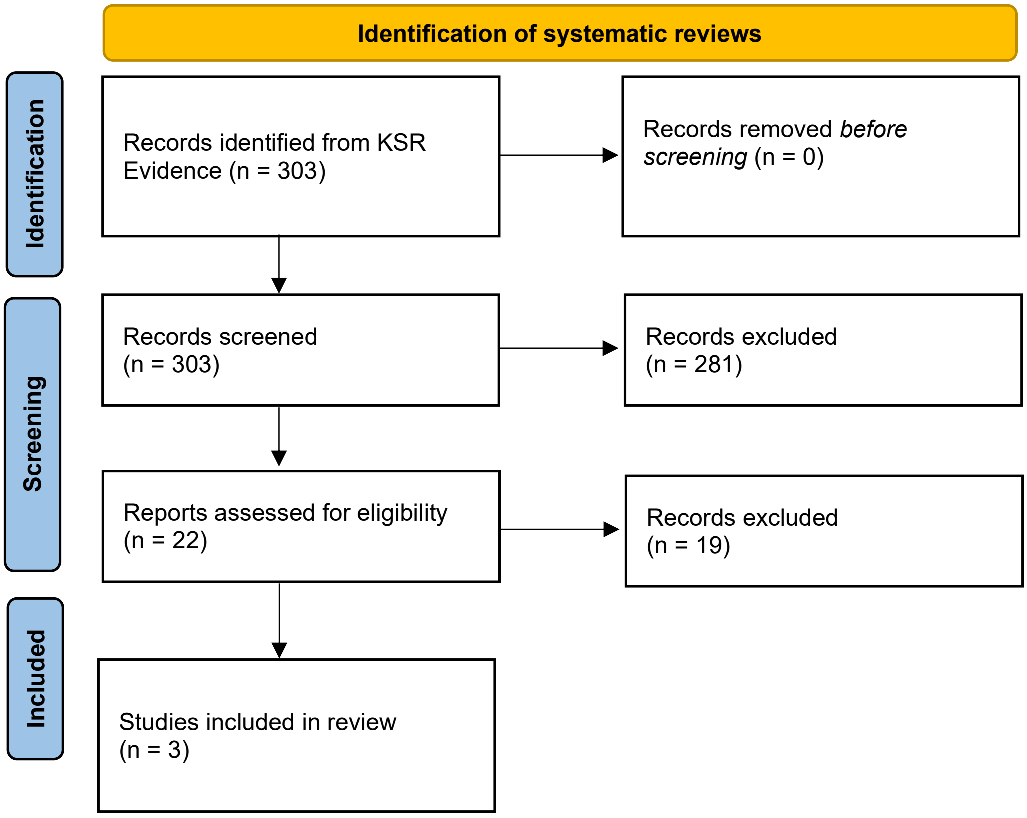

#### Figure S4. Rapid review 4: Combined search of complications and abnormal test results in hypertensive patients

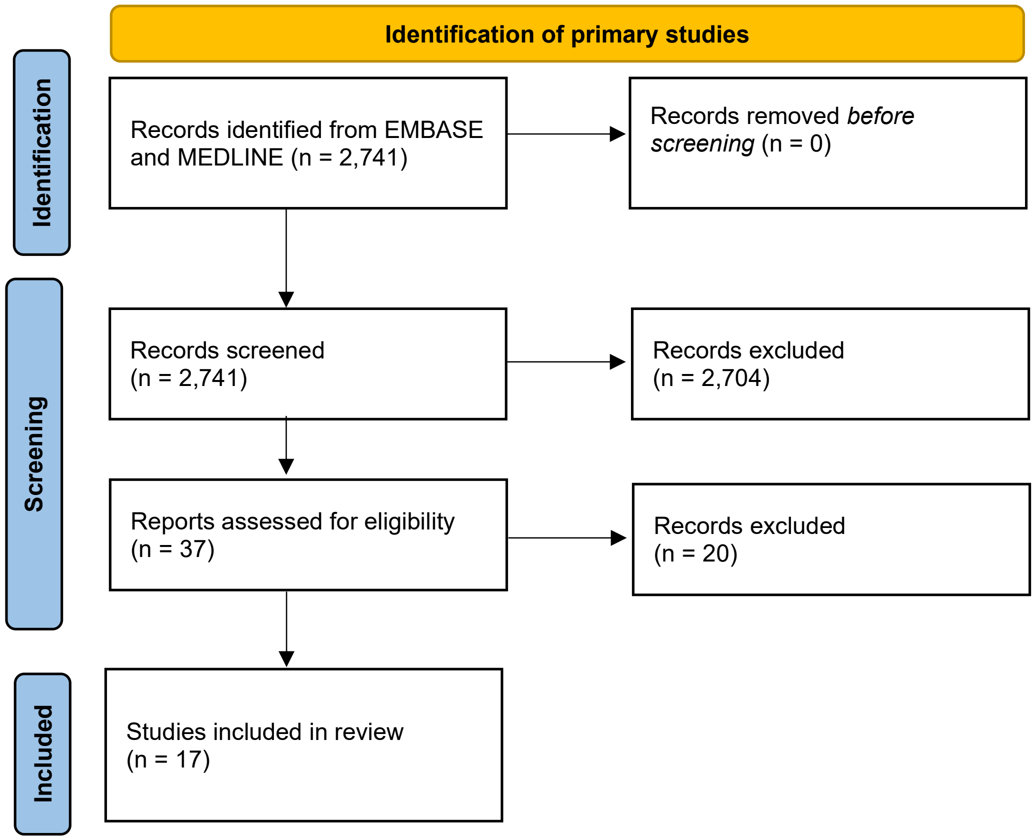


#### Figure S5. Rapid review 5: Hypertension drug side effects

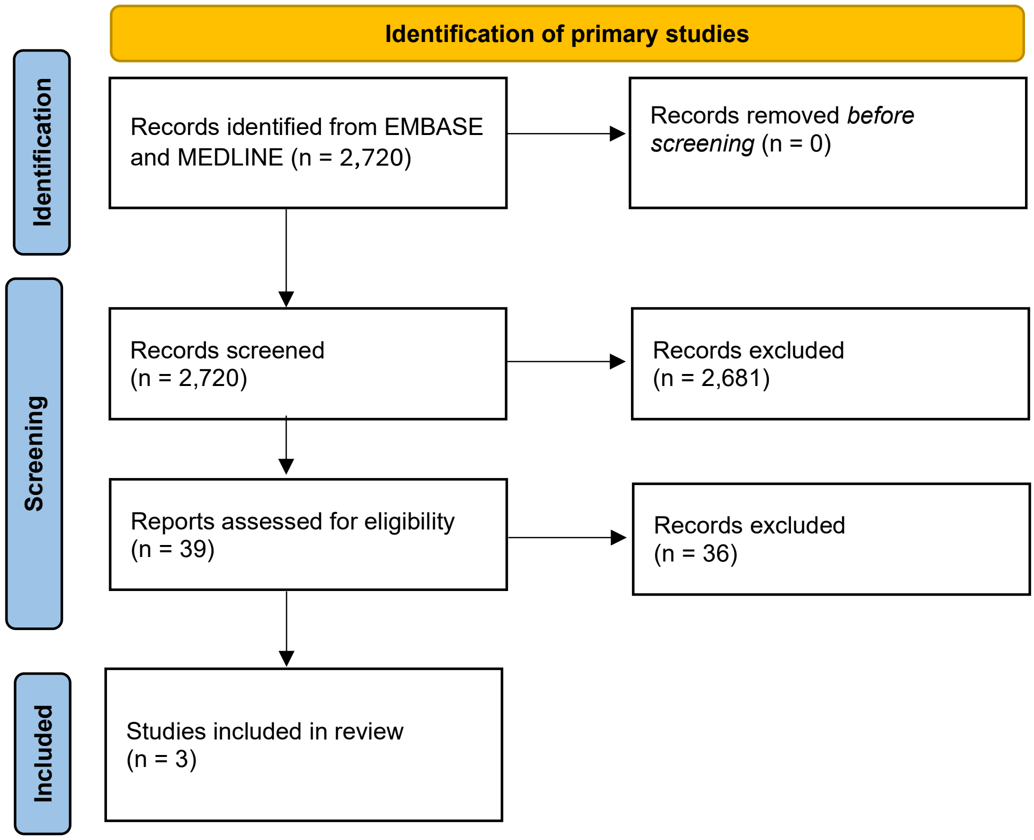
